# Systematic variant-to-gene mapping highlights *TGFB2* and *VEGFA* as adipokine-coding genes with non-obese, insulin-resistance-like characteristics and distinct disease risks

**DOI:** 10.64898/2026.05.01.26352257

**Authors:** Chen-Yang Su, Masashi Hasebe, Adriaan van der Graaf, Yefeng Yang, Hsuan Megan Tsao, Leighton Smith, Guillaume Butler-Laporte, Sirui Zhou, Wenmin Zhang, Tianyuan Lu, Satoshi Yoshiji

## Abstract

Adipokines are key metabolic hormones that modulate cardiometabolic risk through multiple distinct biological pathways. To delineate these pathways, we systematically mapped adipokine⍰associated variants to putative effector genes (V2G) across□1,669 human traits in three ancestries from the Million Veteran Program. Grouping the variants by their associations with insulin⍰resistance-related traits yielded six discrete variant clusters, including a “Lipodystrophy” cluster characterised by lower body⍰mass index but higher waist⍰to⍰hip ratio, fasting glucose, and insulin levels. V2G mapping implicated *TGFB2* and *VEGFA* as candidate effector genes in the Lipodystrophy cluster. *VEGFA* also appeared in a distinct “Thyroid-adiposity” cluster that was strongly associated with increased insulin resistance and decreased thyroid function. The Thyroid-adiposity cluster comprised variants that are thyroid eQTLs, unlike those in the Lipodystrophy cluster. These findings indicate that *VEGFA* may influence insulin resistance via two separate mechanisms: abnormal adiposity and altered thyroid function. Although both clusters increased coronary artery disease risk, only the Lipodystrophy cluster increased type□2 diabetes risk. Our results highlight mechanistically distinct routes by which adipokines modulate insulin resistance and cardiometabolic disease.

## Background

Adipokines are bioactive peptides secreted by adipose tissue that play critical roles in systemic metabolism by regulating insulin sensitivity^1–4^. Dysregulated adipokine secretion is implicated in obesity, type 2 diabetes (T2D), and cardiovascular disease^5–11^. Given their presence in systemic circulation, adipokines represent accessible targets for therapeutic modulation, making them promising drug targets^12–14^. However, despite their central role in metabolic physiology, the genetic and regulatory mechanisms underlying adipokine function in the pathogenesis of metabolic diseases remain incompletely understood.

Genome-wide association studies (GWAS) have identified thousands of loci influencing adiposity and cardiometabolic traits, yet translating these signals into causal variants, genes, and mechanisms remains a major challenge. The vast majority of trait-associated variants reside in non-coding regions, complicating efforts to identify the effector genes and their tissue-or cell-type-specific functions^15^. As a result, many GWAS loci lack clear biological interpretation, limiting the ability to translate genetic findings into mechanistic insights. One strategy to address these barriers is variant-to-gene (V2G) mapping, which aims to link genetic variants to their effector genes. Integrative V2G approaches, such as the combined SNP-to-gene (cS2G) framework^16^, aggregate evidence from multiple sources including functional annotations, expression quantitative trait loci (eQTL), enhancer-gene interactions, and chromatin contact maps to systematically prioritize likely effector genes. This framework elucidates the biological roles of variants by linking them to their most likely effector genes, including those involved in adipokine signaling.

T2D and insulin resistance (IR) are genetically heterogeneous disorders. Studies have clustered T2D^17^ and fasting insulin loci^18^ based on their associations with metabolic traits and identified distinct mechanistic clusters reflecting biologically meaningful pathways. Such clustering of variants based on biomarkers or disease traits has important implications. First, clustering can improve interpretation of genetic variants, particularly those located in non-coding regions with ambiguous functions, by highlighting biologically relevant groupings. Second, when clustering incorporates disease-associated traits, it can further refine variant classification by prioritizing variants implicated in specific pathogenic mechanisms. Given the established connection between adipokines and metabolic physiology, clustering variants mapped to adipokine-related genes offers a promising yet underexplored avenue to dissect the mechanisms underlying metabolic disorders.

Here, we conducted systematic variant-to-gene mapping and multi-ancestry fine-mapping across 1,669 traits in the Million Veteran Program (MVP)^19^ to link fine-mapped variants to putative effector adipokine genes. Using a clustering approach, we identified mechanistically distinct groups of adipokine-linked variants, characterized by shared IR-trait signatures within each group, highlighting key genes such as *VEGFA* and *TGFB2.* We integrated multiple lines of genetic and epigenomic evidence to validate these clusters. Our findings offer insights into the heterogeneity of IR and advance understanding of how adipose-related variants influence cardiometabolic disorders, establishing a framework for dissecting the mechanistically distinct routes through which adipokines modulate disease risk.

## Results

An overview of this study is shown in **Figure 1**. Briefly, we began with GWAS from MVP (∼20 million variants) and subsetted to variants with a combined SNP-to-gene (cS2G) annotation/linking score (3,510,384 variants), which integrates multiple sources of SNP-to-gene evidence (e.g., coding and promoter annotation, fine-mapped cis-eQTLs, and enhancer–gene/chromatin interaction links) to nominate putative effector genes for each variant then performed multi-ancestry fine-mapping with SuSiEx for each trait across the biobank. We then assigned a representative effector gene to each credible set by summing variant-level posterior inclusion probabilities (PIPs) across all variants linked by cS2G to the same gene within that credible set and selecting the gene with the largest summed PIP. If a variant was linked to multiple genes by cS2G, its full PIP contributed to each linked gene. Finally, we investigated adipokine genes and performed hierarchical clustering of variants to identify distinct genetic clusters and pinpointed a Lipodystrophy cluster and a Thyroid-adiposity cluster with distinct cardiometabolic trait signatures.

**Figure 1.**
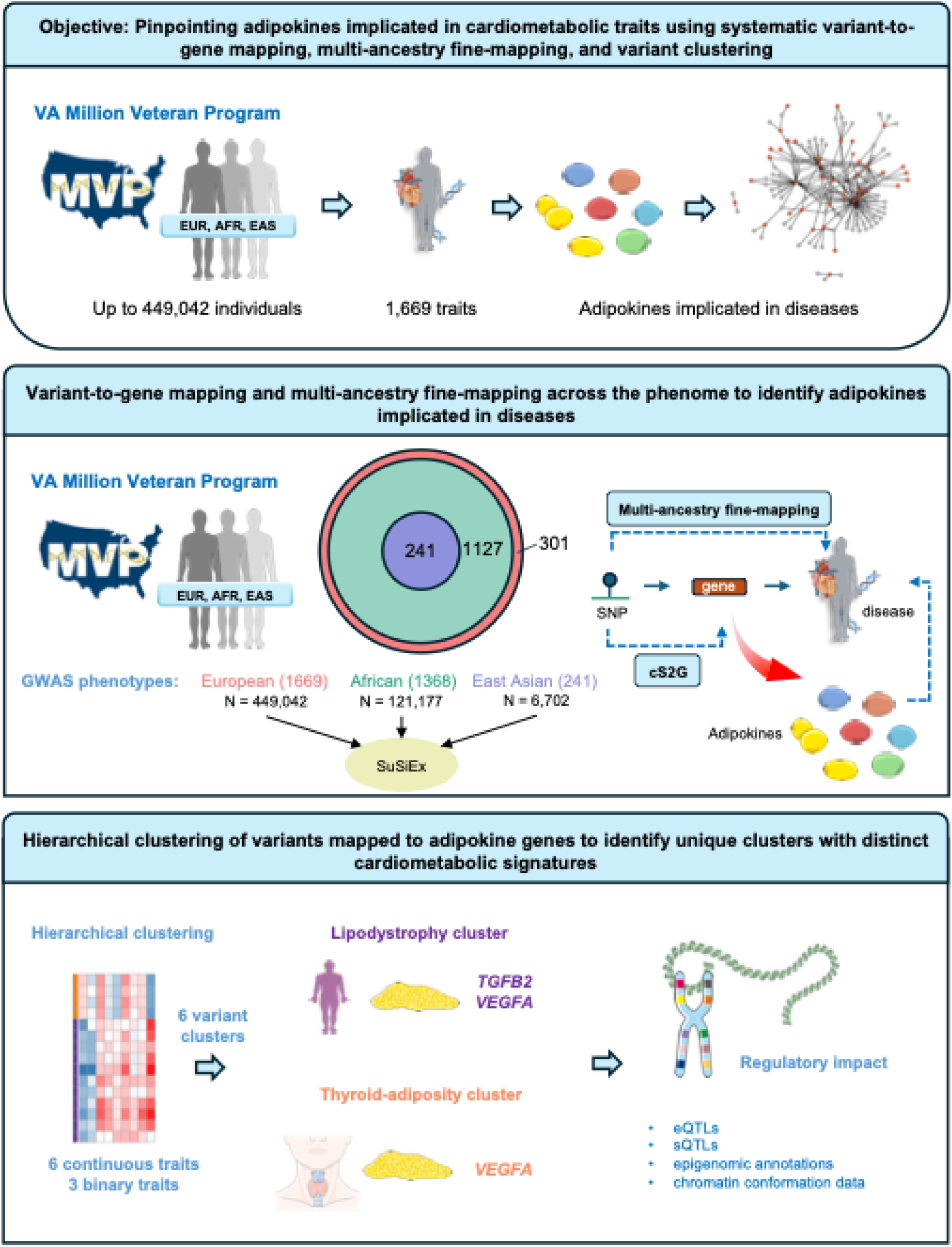
Study design. In this study, we analyzed publicly available genome-wide association study (GWAS) data from the Million Veteran Program (MVP) spanning European (EUR; *n* = 449,042), African (AFR; *n* = 121,177), and East Asian (EAS; *n* = 6,702) ancestries, assessing 1,669, 1,368, and 241 traits, respectively. We implemented the combined SNP-to-gene (cS2G) strategy to annotate genetic variants, identifying 3,510,384 variants with cS2G annotation scores. To identify likely causal variants, we conducted cross-ancestry fine-mapping using SuSiEx and pinpointed variants with adipokine-coding genes as putative effector genes. Hierarchical clustering was performed on these adipokine gene variants and showed six distinct clusters including a Lipodystrophy cluster and a Thyroid-adiposity cluster. These two clusters were further explored for their regulatory impact through expression quantitative trait loci, epigenomic annotations, and chromatin conformation data. GWAS: genome-wide association study; cS2G: combined SNP-to-gene; eQTL: expression quantitative trait loci; sQTL: splicing quantitative trait loci.

### Variant-to-gene and multi-ancestry fine-mapping

We used GWAS from the MVP for traits measured in European (EUR; *n* = 449,042 individuals; 1,669 traits), African (AFR; *n* = 121,177 individuals; 1,368 traits), and East Asian (EAS; *n* = 6,702 individuals; 241 traits) ancestries to perform V2G mapping and multi-ancestry fine-mapping, aiming to identify variants with adipokine putative effector genes associated with diseases (**Supplementary Table 1-3**, **Figure 1**, and **Supplementary Note 1**). For V2G mapping, we applied cS2G to assign SNP-gene linking scores genome-wide, thereby prioritizing putative effector genes based on integrated functional genomic evidence rather than proximity alone. After mapping association results to 3,510,384 variants with a cS2G annotation score (**Methods**), we performed fine-mapping across up to three ancestries with SuSiEx^20^, a fine-mapping method that leverages cross-ancestry differences in linkage disequilibrium (LD) patterns to improve resolution (**Supplementary Note 2** and **3** and **Methods**).

### Clustering of genetic variants identifies distinct insulin-resistance like groups acting through distinct mechanisms

By linking these genetic variants to candidate genes and refining causal signals through fine-mapping, we gained insight into how specific variants contribute to disease. We compiled a curated list of adipokine-coding genes from published catalogs and targeted literature searches, selecting 90 genes with prior evidence of adipokine activity (**Supplementary Table 4**). Using this list, we identified 738 gene-trait pairs (515 unique) involving 52 genes across 203 traits (**Supplementary Table 5**). After additional trait filtering (see **Methods**), 40 adipokine genes remained associated with 107 traits, comprising 228 unique connections (**Figure 2**). Examination of SNP-gene-trait triplets revealed 163 unique variants with adipokine-coding genes as putative effector genes. From here on, we refer to these as adipokine gene variants for simplicity (e.g., *VEGFA* variants). These gene-trait associations included well-known examples, such as *LPL* with triglycerides (TG), high-density lipoprotein cholesterol (HDL-C), and low-density lipoprotein cholesterol (LDL-C), as well as emerging targets such as *CXCL12* with myocardial infarction^21^ and *TGFB2* and *VEGFA* with T2D^22,23^.

**Figure 2.**
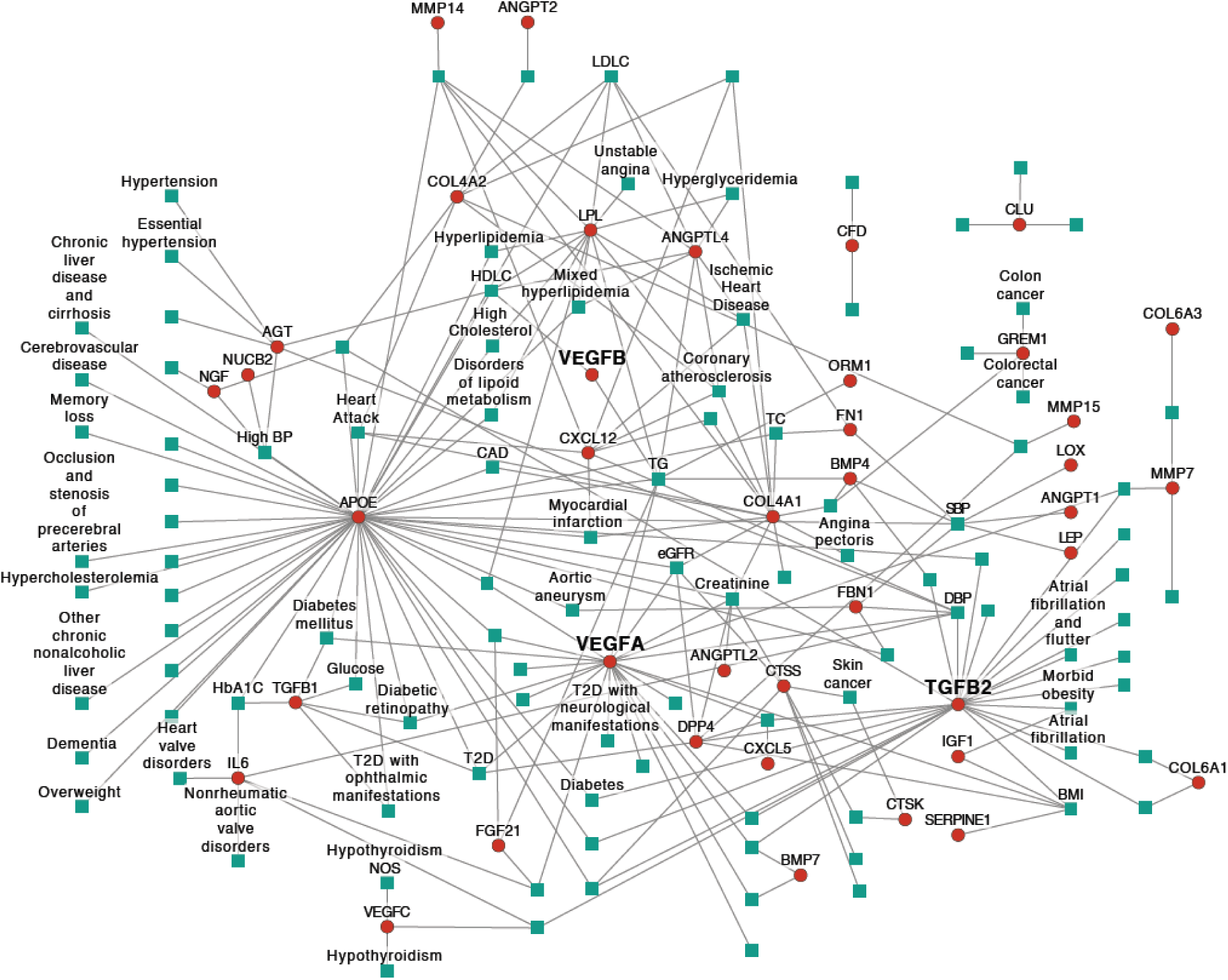
Adipokine gene-trait network. Results from combined SNP-to-gene (cS2G) and multi-ancestry fine-mapping that link adipokine genes to traits are shown. For simplicity, only adipokine genes are displayed. The plot depicts 40 genes connected to 107 traits with 228 unique connections. Proteins are represented by red circles, while outcomes are presented as teal squares. Proteins and outcomes are connected by gray edges. Only relevant outcomes are labeled to simplify visualization. The font size for *VEGFA*, *VEGFB*, and *TGFB2* is increased for visibility.

Since IR is closely associated with altered adipokine secretion and contributes to metabolic dysfunction, we further characterized these variants specifically in the context of IR. We used summary statistics from external sources spanning adiposity and fat distribution (body mass index [BMI], body fat percentage [BFP], waist-hip-ratio adjusted for body mass index [WHRadjBMI], and visceral-to-gluteofemoral adipose tissue ratio [VATtoGFAT]), glycemic traits (HbA1c, fasting glucose, and fasting insulin), and cardiometabolic outcomes (T2D, coronary artery disease [CAD], and ischemic stroke), with sample sizes ranging from 39,076 to 1,812,017 (European ancestry) (**Supplementary Table 6**) and performed hierarchical clustering based on trait associations. We identified six distinctive clusters of genetic variants with characteristic patterns of trait similarity (**Supplementary Figure 1;** see **Methods**). We aligned all variants such that the effect estimates correspond to the alleles associated with increased WHRadjBMI. Distinct patterns of the clusters are shown in **Figure 3a**, involving 156 SNPs that were found in the WHRadjBMI GWAS. Based on their multi-trait patterns, we labeled the six clusters as follows: Cluster 1, Residual effects; Cluster 2, Thyroid-adiposity; Cluster3, Lipodystrophy; Cluster 4, CAD-increasing; Cluster 5, CAD-decreasing; and Cluster 6, APOE.

**Figure 3.**
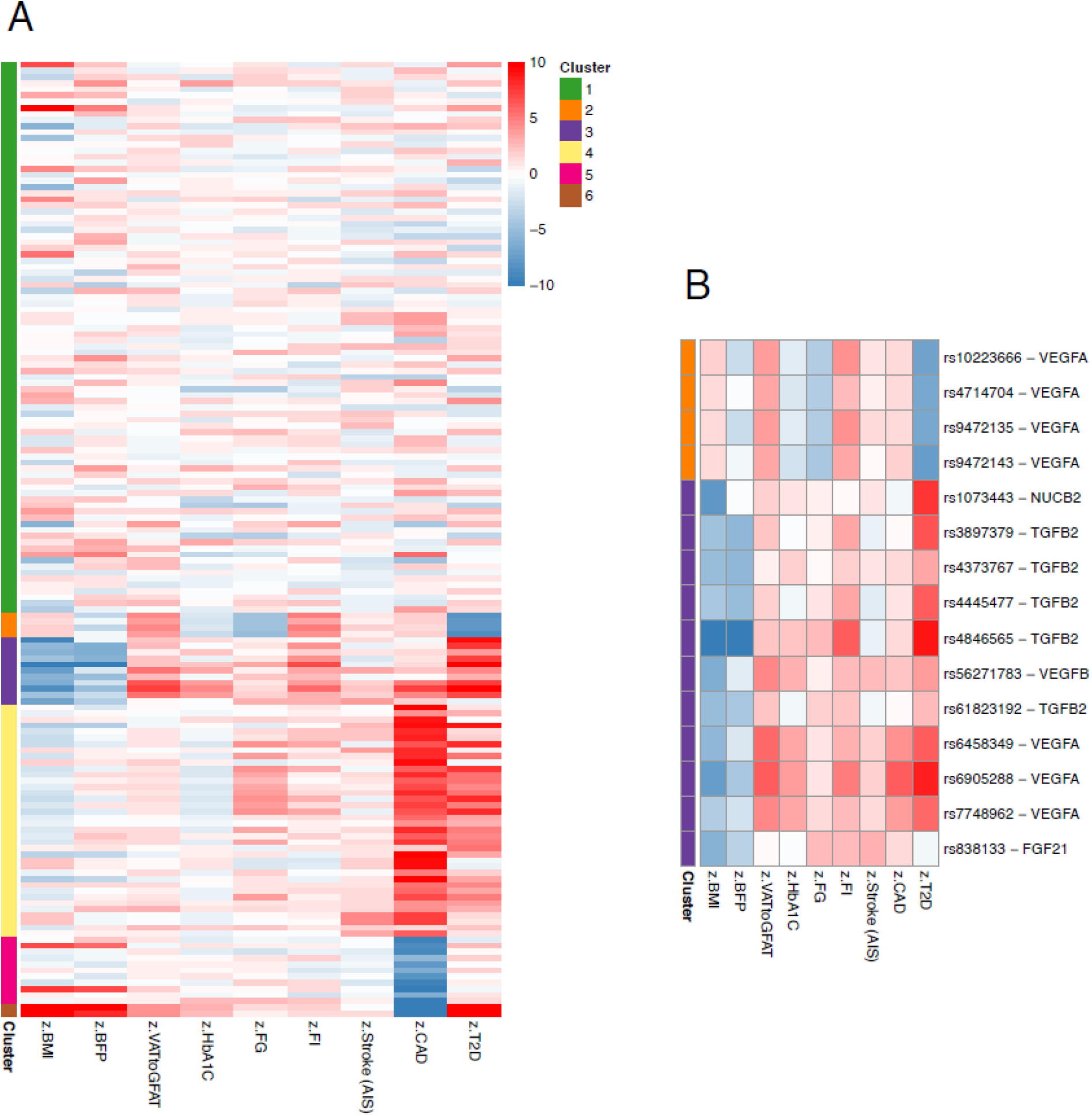
Clustering of genetic variants. (a) Heatmap of hierarchical clustering of genetic variants. Using biomarker and cardiovascular binary disease traits, we created a heatmap of Z scores for top posterior inclusion probability (PIP)-scoring variants in adipokine gene-trait pairs. The matrix of Z scores from GWAS of six continuous traits (body mass index (BMI), body fat percentage (BFP), ratio of visceral to gluteofemoral adipose tissue (VATtoGFAT), glycated hemoglobin (HbA1c), fasting glucose (FG), and fasting insulin (FI)) and three disease traits (stroke (any ischemic stroke; AIS), coronary artery disease (CAD), and type 2 diabetes (T2D)) is shown. All variants have been aligned to increase waist-to-hip ratio adjusted for BMI (WHRadjBMI). (b) Zoomed-in view of Cluster 2 (orange) and Cluster 3 (purple). Red indicates that the variant is associated with an increase in the trait displayed on the x-axis, whereas blue indicates a decrease. The scale ranges from -10 < *Z* < 10. The y-axis lists variants mapped to genes identified by cS2G and multi-ancestry fine-mapping.

Variants in each cluster showed distinct association patterns with biomarkers and diseases. Cluster 1 comprised variants that showed relatively modest and heterogeneous associations with these traits and was considered a residual cluster. Cluster 2 was characterized by variants associated with increased BMI, insulin, visceral adiposity, and increased risk for cardiovascular disease, but lower risk of T2D, recapitulating non-diabetogenic hyperinsulinemia as previously reported^18^. Cluster 3 showed a distinct lipodystrophy-like signature, with variants associated with lower BMI and BFP but higher levels of glycaemic biomarkers and increased cardiometabolic risk^17,18,24–27^. Notably, these variants were strongly associated with T2D (median *Z* = 7.2) compared to with CAD (median *Z* = 1.6). Cluster 4 was characterized by variants showing strong positive associations with CAD but weak associations with T2D, whereas Cluster 5 showed negative associations with CAD risk. Variants mapped to *APOE* were found in Clusters 1, 4, 5, and 6, demonstrating the highly pleiotropic nature of this locus. Interestingly, variants mapped to *LPL* were found in Clusters 1 and 4 only, but not in Cluster 3, suggesting that these variants may contribute to increased CAD risk.

**Figure 3b** illustrates the “Thyroid-adiposity cluster” (Cluster 2) and “Lipodystrophy cluster” (Cluster 3) in greater detail. Cluster 3 exhibited a distinct metabolic profile characterized by low BMI (*Z* < -4.7), low BFP (*Z* < -0.45) but generally high ratio of visceral (VAT) to gluteofemoral (GFAT) adipose tissue (VATtoGFAT), glycated hemoglobin (HbA1c), fasting glucose, fasting insulin as well as increased risk of stroke (any ischemic stroke; AIS), CAD, and T2D, indicative of a clear lipodystrophy-like pattern (**Figure 3b**). We therefore refer to this as a “Lipodystrophy cluster”. This lipodystrophy-like, or non-obese, IR cluster included variants mapped to five genes including *TGFB2*, *VEGFA*, *VEGFB*, *NUCB2*, and *FGF21*. For *VEGFB* (rs56271783), *NUCB2* (rs1073443), and *FGF21* (rs838133), fine-mapping resolved a single likely causal variant for each gene. All three genes have been previously reported in association with metabolic abnormalities, including obesity and other adverse metabolic phenotypes^18,28,29^. Notably, four variants mapped to *VEGFA*, which encodes vascular endothelial growth factor A, and were grouped into Cluster 2, while 11 variants mapping to *VEGFA*, *TGFB2*, *NUCB2*, *VEGFB*, and *FGF21* were grouped into Cluster 3. Further details of these two clusters are provided below.

### Variant clustering identifies a lipodystrophy-like insulin resistance cluster

The five variants in the Lipodystrophy cluster that mapped to *TGFB2* were located in *trans* spanning a ∼465kb region distal to the *TGFB2* gene, which encodes transforming growth factor beta-2 (TGFβ-2), a key protein regulating angiogenesis, muscle tissue regulation, and body fat development^30^. The farthest (rs3897379; chr1:219586391; GRCh38) and nearest (rs61823192; chr1:219121228; GRCh38) variants lay ∼1.24Mb and ∼0.78Mb downstream of the *TGFB2* transcription start site, respectively. rs4846565 has previously been reported as a locus influencing glycemic traits related to insulin metabolism and regulation^31–33^. In our analysis, we fine-mapped this variant as a likely causal signal for several metabolic traits (**Supplementary Table 5**), and phenome-wide association results were also concordant (**Supplementary Figure 2**). Ancestry-specific support for the highlighted TGFB2 signals is summarized in **Supplementary Table 7**. Across the TGFB2 highlighted variant-trait credible-sets, all had EUR post-hoc probability >0.8, and the subset with AFR post-hoc probability >0.8 showed concordant EUR/AFR effect directions; more broadly, available EUR and AFR effect estimates were directionally concordant where both were present. EAS support was limited, consistent with the smaller MVP East Asian sample size and trait coverage.

#### Regulatory role of lipodystrophy-like insulin resistance genetic variants linked to TGFB2

We next aimed to determine whether *TGFB2* variants in the Lipodystrophy cluster have regulatory potential in adipose tissues and related cell types. We assessed chromatin accessibility from ENCODE^34^ and RegulomeDB^35^, and examined physical interactions with promoter regions using promoter Capture Hi-C (pCHi-C)^36^ data from preadipocytes^37^. Notably, the rs61823192-*TGFB2* link was supported by multiple lines of epigenomic evidence (**Figure 4**). pCHi-C data in preadipocytes revealed looping interactions between this locus and the promoter region of *TGFB2* (**Figure 4a**). ATAC-seq data indicated that the variant was located in regions of open chromatin in subcutaneous adipose tissue and mesenteric fat. The variant also overlapped active histone marks (H3K27ac in subcutaneous abdominal adipose tissue and H3K4me3 in adipocytes), consistent with enhancer or promoter activity (**Figure 4b**). Phenome-wide association analyses showed positive associations of rs61823192 (chr1:219121228 -T) with WHRadjBMI (β = 0.042, *P* = 2.07 × 10^-11^) and triglycerides (β = 0.034, *P* = 2.84 × 10^-12^) (**Figure 4c**). Together, these findings suggested that rs61823192 may act as a regulatory variant that modulates *TGFB2* expression and adipose tissue function.

**Figure 4.**
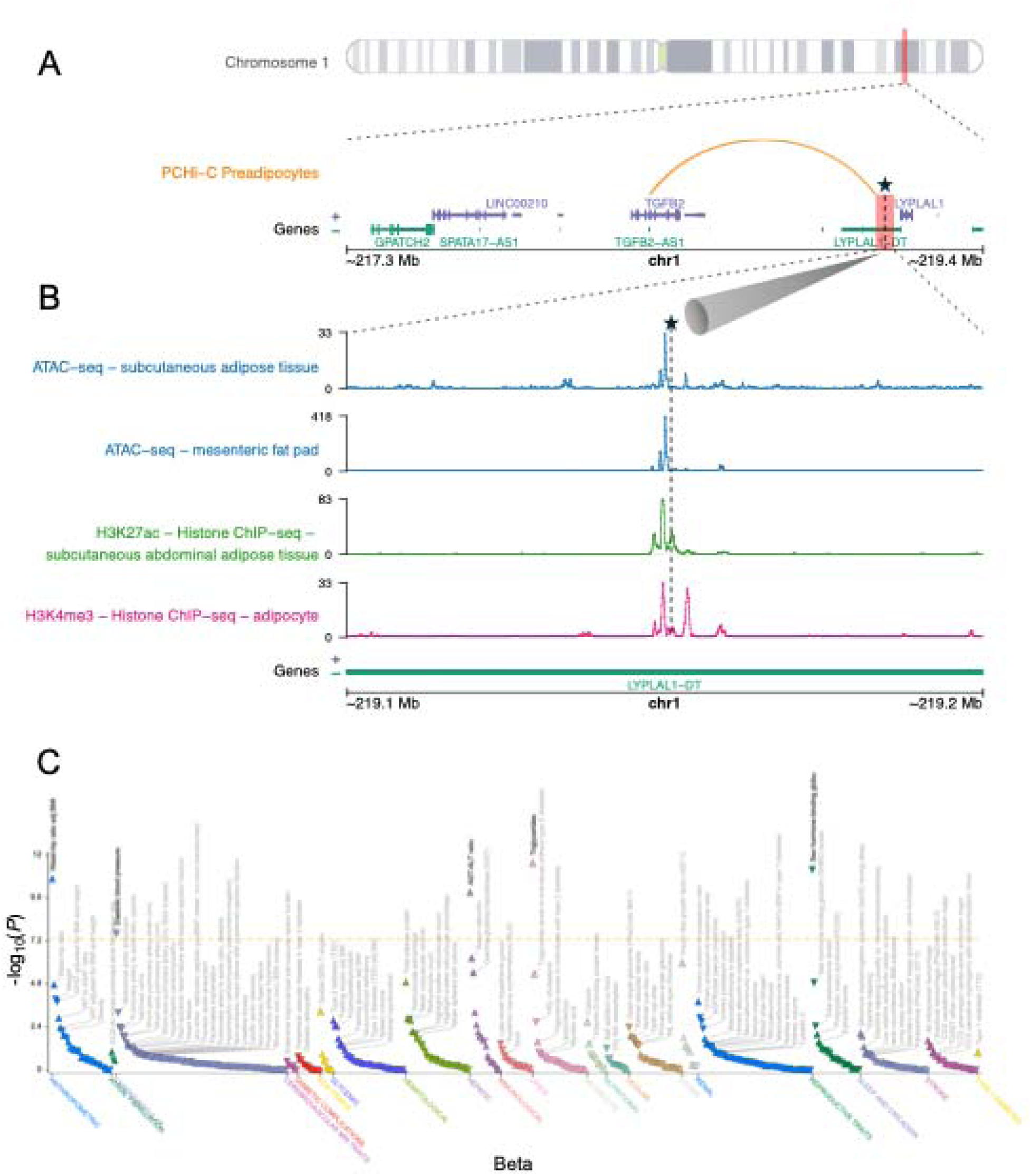
Regulatory evidence and PheWAS for rs61823192 mapped to *TGFB2* in the Lipodystrophy cluster. (a) Cytogenetic location of the *TGFB2* gene. The red-shaded region (chr1:217345336—219444619; GRCh38) represents a ±1 Mb window surrounding the *TGFB2* gene, shown at higher resolution with the gene track. The arc depicts promoter capture Hi-C (PCHi-C) results from preadipocytes Garske et al^37^. The black star denotes rs61823192, and the dashed connector lines indicate the genomic region shown at higher resolution and carried forward to panel (b). (b) Epigenomic markers within a ±30 kb window around rs61823192 (chr1:219121228; GRCh38). The top four panels show ATAC-seq (blue), H3K27ac ChIP–seq (green), and H3K4me3 ChIP–seq (pink) data from relevant tissues. These datasets are publicly available through ENCODE^34^ and RegulomeDB^35^ (ENCODE accession IDs: ENCFF241AEA, ENCFF565QME, ENCFF080SAC, and ENCFF573BYG). The bottom panel displays gene tracks. The vertical dashed line (and star) marks the position of rs61823192 across all tracks. (c) Phenome-wide association study (PheWAS) results for rs61823192. Triangles indicate the direction of effect for the variant: upward triangles denote a positive effect estimate of the T allele, and downward triangles denote a negative effect estimate.

### Identification of a Thyroid-adiposity cluster

Within Cluster 2, we found four *VEGFA* variants (rs10223666, rs4714704, rs9472135, rs9472143) (**Figure 3b**). These four variants had WHRadjBMI-increasing alleles that were also associated with greater BMI (*Z* > 1.5), VATtoGFAT (*Z* > 4.0), and fasting insulin (*Z* > 3.3), while associating with lower glycemic trait levels (fasting glucose, *Z* < -4.6; HbA1c, *Z* < -1.7). Notably, these alleles were strongly associated with a decreased risk of T2D (*Z* < -7.7) and showed marginally increased risks of stroke (*Z* > 0.42) and CAD (*Z* > 1.6). Taken together, these features suggested an obesity-like IR cluster characterized by non-diabetogenic hyperinsulinemia and lower glycemic trait levels, recapitulating the previously described “Preserved Insulin Secretion (PIS)” cluster by Sevilla-González et al^18^. Ancestry-specific support for the highlighted *VEGFA* signals was strongest in European ancestry and remained directionally concordant with African ancestry estimates with limited support in East Asian MVP analyses due to limits in sample size and trait coverage in this ancestry (**Supplementary Table 7**).

Most phenotype-associated variants are located in non-coding regulatory regions, and their mechanisms often remain unclear. Consistent with this, all four *VEGFA* variants were intronic. To evaluate potential regulatory effects, we assessed tissue-specific gene expression, epigenomic annotations, and phenome-wide association studies.

#### Gene expression and alternative splicing patterns in different tissues

To determine tissue specificity, we queried GTEx v8^38^. All four variants were *VEGFA* expression quantitative trait loci (eQTLs) in thyroid tissue (*P* < 1.2 × 10^-4^) (**Supplementary Table 8**). The variants were also significantly associated with expression changes in long intergenic non-protein coding RNA (**Supplementary Note 4**). To gain further insights, we queried them in AdipoExpress^39^, an eQTL meta-analysis of 2,344 subcutaneous adipose tissue samples, and observed significant associations for all four variants with *VEGFA* expression (*P* < 0.028) (**Supplementary Table 9**), supporting regulatory effects relevant to adipose biology (**Supplementary Figure 3-6**).

#### Epigenomic evidence

Next, to better understand the potential regulatory roles of the identified intronic *VEGFA* variants, we searched for epigenomic evidence using RegulomeDB^35^, which integrates diverse functional genomic datasets to predict regulatory functionality. Variant rs9472135 received a RegulomeDB rank of 1b, indicating strong regulatory potential. This rank reflects the presence of multiple regulatory features, including eQTL and chromatin accessibility QTL (caQTL), transcription factor (TF) binding, the presence of a known motif, DNase footprinting, and a chromatin accessibility peak. In contrast, the other three intronic *VEGFA* variants (rs10223666, rs4714704, and rs9472143) were assigned a rank of 1f, suggesting weaker but still notable regulatory evidence. Each of these variants had a reported score of 0.55436, whereas rs9472135 had 0.76, highlighting rs9472135 as the strongest epigenomic candidate among the four.

#### Phenome-wide association analyses

In phenome-wide association analyses, all four *VEGFA* variants showed their most significant associations with higher thyroid-stimulating hormone (TSH) (*n* = 635,908) and lower free thyroxine (FT4) levels (*n* = 117,166), a combination characteristic of overt (primary) hypothyroidism^40^ (**Supplementary Figure 7-10**). For instance, rs10223666 (chr6:43837765 -C [effect allele]) was associated with increased TSH (β = 0.096, *P* = 5.7 × 10^-226^) and decreased FT4 (β = -0.032, *P* = 1.1e-11). This profile is consistent with the classical negative feedback loop in thyroid regulation, where reduced circulating FT4 alleviates feedback inhibition on the pituitary and is accompanied by elevated TSH^41^. *VEGFA* encodes a potent angiogenic factor important in thyroid physiology; notably TSH stimulation directly upregulates *VEGFA* expression in thyrocytes to promote angiogenesis and glandular hypervascularity^42^. Consistent with these mechanisms, *VEGFA* enhancer polymorphisms that alter transcription factor binding and *VEGFA* expression have been linked to inter-individual differences in TSH and FT4 levels^43^.

Altogether, these lines of evidence supported designation of this group as the “Thyroid-adiposity cluster”.

### Dissecting the regulatory complexity at the VEGFA locus reveals evidence of functional pleiotropy

We next distinguished *VEGFA* variants identified in Cluster 2 from those in Cluster 3 to assess the target genes and tissues in which each cluster exerts its effects. The three *VEGFA* variants in Cluster 3 (rs6458349, rs6905288, rs7748962) showed no *VEGFA* eQTL evidence in GTEx v8 in any tissue, yet all three were associated with *VEGFA* expression in subcutaneous adipose tissue in AdipoExpress^39^ (*P* < 0.05) (**Supplementary Table 9** and **Supplementary Figure 11-13**). Specifically, rs6905288 (chr6:43791136-A) showed colocalization between GWAS signals for CAD (colocalization probability = 0.99), cholesterol (colocalization probability = 0.83), and WHRadjBMI (colocalization probability = 1.0) and its corresponding eQTL in adipose tissue, suggesting involvement of *VEGFA* in these metabolic-related traits (**Supplementary Table 10**). In contrast to the Cluster 2 *VEGFA* variants, we found no evidence of association of the Cluster 3 variants with gene expression in thyroid tissues or with thyroid-related traits through phenome-wide association analyses (**Supplementary Figure 14-16**). These results support a distinct mechanism for the Cluster 3 *VEGFA* variants from that implicated in Cluster 2 (**Supplementary Note 5**).

### Circulating VEGFA and TGF-β2 levels and metabolic outcomes in the UK Biobank

Given the observed genetic and regulatory links of *VEGFA* and *TGFB2* with metabolic traits, we next investigated how their corresponding proteins were related to metabolic traits and disease outcomes in the UK Biobank, a large population-based clinical cohort. We analyzed cross-sectional associations between baseline circulating VEGFA and TGF-β2 levels and adiposity-related traits in up to 44,000 participants (**Table 1**). Higher VEGFA levels were strongly associated with greater BMI (β = 0.17, 95% CI 0.16–0.18, *P* = 2.4 × 10^-287^), WHRadjBMI (β = 0.061, 95% CI 0.055–0.067, P = 5.0 × 10^-85^), and TG to HDL-C ratio (β = 0.20, 95% CI 0.19–0.21, P < 1.0 × 10^-300^) (**Figure 5A**). For TGF-β2, associations were weaker but directionally consistent (**Figure 5B**). Higher TGF-β2 was associated with greater BMI (β = 0.031, 95% CI 0.022–0.040, P = 6.5 × 10^-11^), WHRadjBMI (β = 6.9 × 10^-3^, 95% CI 8.9 × 10^-4^–1.3 × 10^-2^, *P* = 0.024), and TG to HDL-C ratio (β = 0.054, 95% CI 0.045–0.064, *P* = 1.2 × 10^-28^). Five of the six associations were significant after Bonferroni correction (*P* < 0.0083), whereas the association between TGF-β2 and WHRadjBMI was nominally significant.

**Figure 5.**
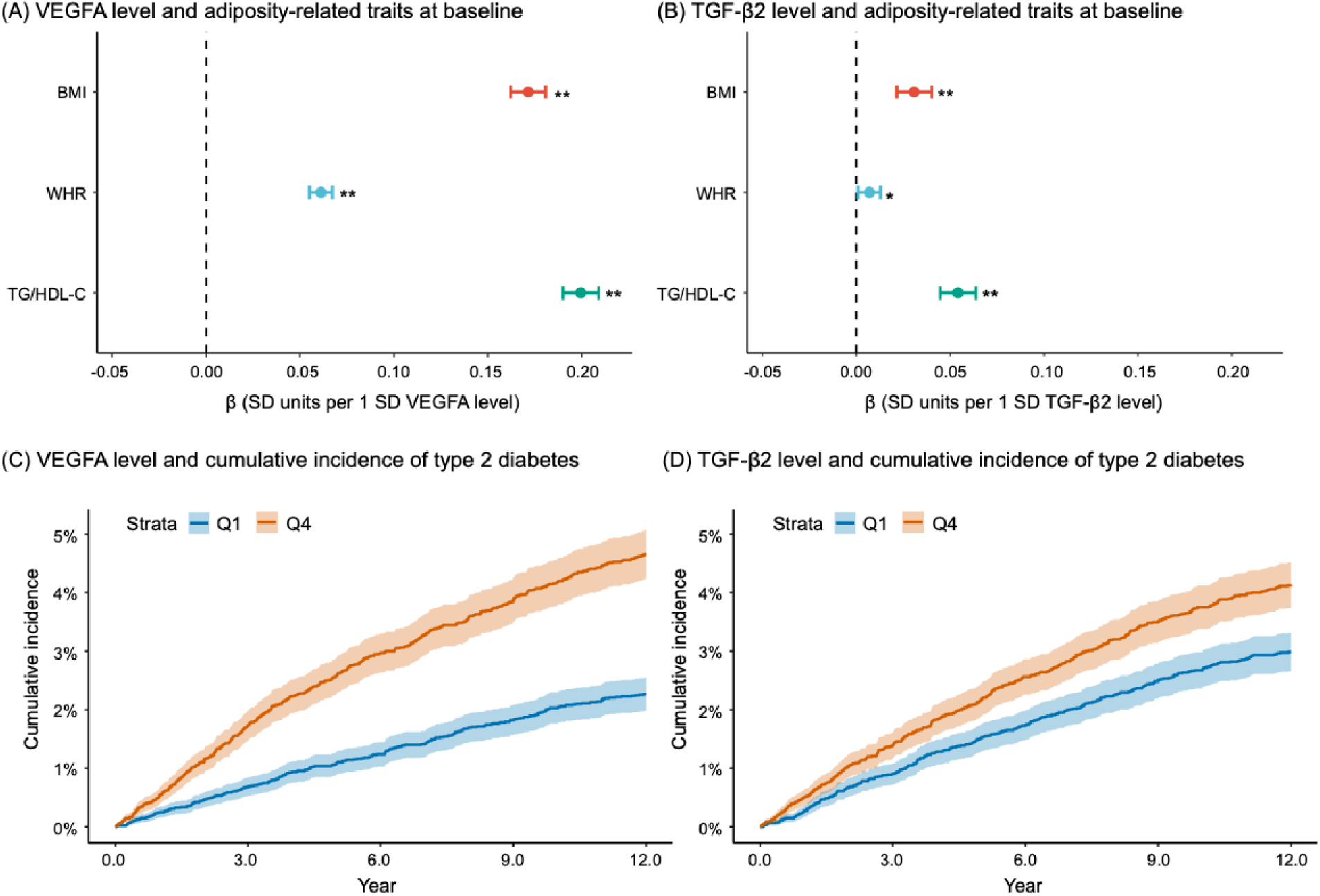
Associations of baseline VEGFA and TGF-β2 levels with adiposity-related traits and incidence of type 2 diabetes in the UK Biobank. **(A, B)** Associations between circulating VEGFA and TGF-β2 levels and adiposity-related traits at baseline were estimated by linear regression, with both the protein (exposure) and each trait (outcome) standardized to mean 0 and SD 1. Points denote standardized β coefficients and bars indicate 95% CIs for body mass index (BMI), waist-to-hip ratio (WHR), and triglycerides (TG) to high-density lipoprotein cholesterol (HDL-C) ratio (TG/HDL-C). β represents the change in the trait (in SD units) per 1-SD higher protein level. In analyses using WHR as the outcome, BMI was included as an additional covariate. \**P* < 0.05, nominal association; \*\**P* < 0.0083 (Bonferroni correction for six tests). **(C, D)** Cumulative incidence of type 2 diabetes according to quartiles of baseline VEGFA and TGF-β2 levels. Curves compare the lowest quartile (Q1, blue) and highest quartile (Q4, orange); shaded bands show 95% CIs.

**Table 1.**
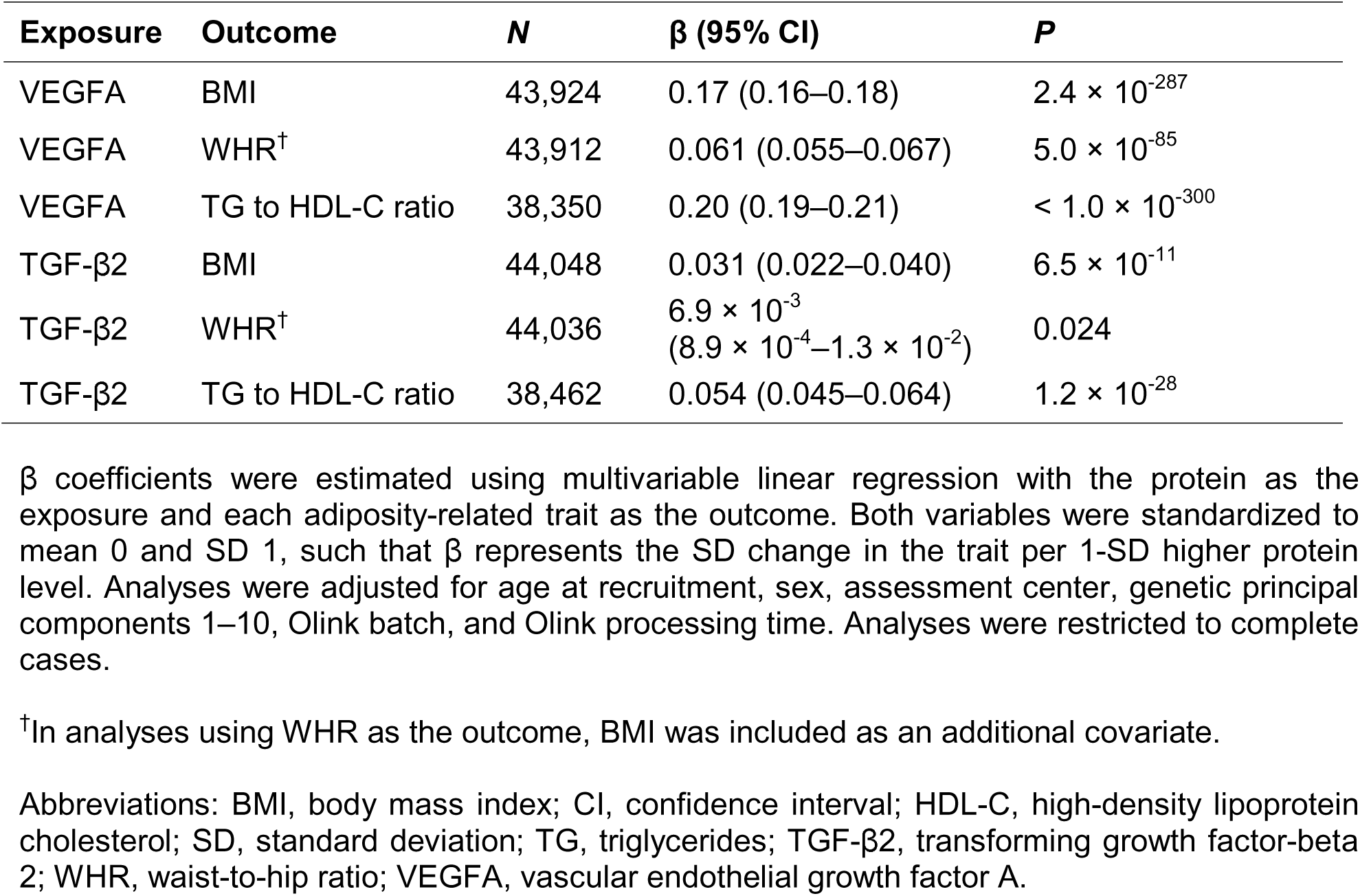
Associations of baseline VEGFA and TGF-β2 levels with adiposity-related traits and incidence of type 2 diabetes in the UK Biobank.

We then examined the prospective associations of circulating VEGFA and TGF-β2 levels with incident T2D over a median follow-up of 13.2 years. For VEGFA, the analysis included 41,779 participants and 1,546 incident T2D cases. For TGF-β2, the analysis included 41,882 participants and 1,561 incident T2D cases. In Cox proportional-hazards regression comparing the highest and lowest quartiles of baseline protein levels (Q4 vs. Q1), higher VEGFA was significantly associated with a nearly twofold higher risk of T2D (HR 1.92, 95% CI 1.64–2.24, *P* = 1.7 × 10^-16^). Higher TGF-β2 levels were also significantly associated with increased risk (HR 1.35, 95% CI 1.17–1.56, *P =* 5.3 × 10^-5^). Detailed results are provided in **Table 2**, and cumulative-incidence curves for Q4 vs. Q1 are shown in **Figure 5C** and **Figure 5D**.

**Table 2.**
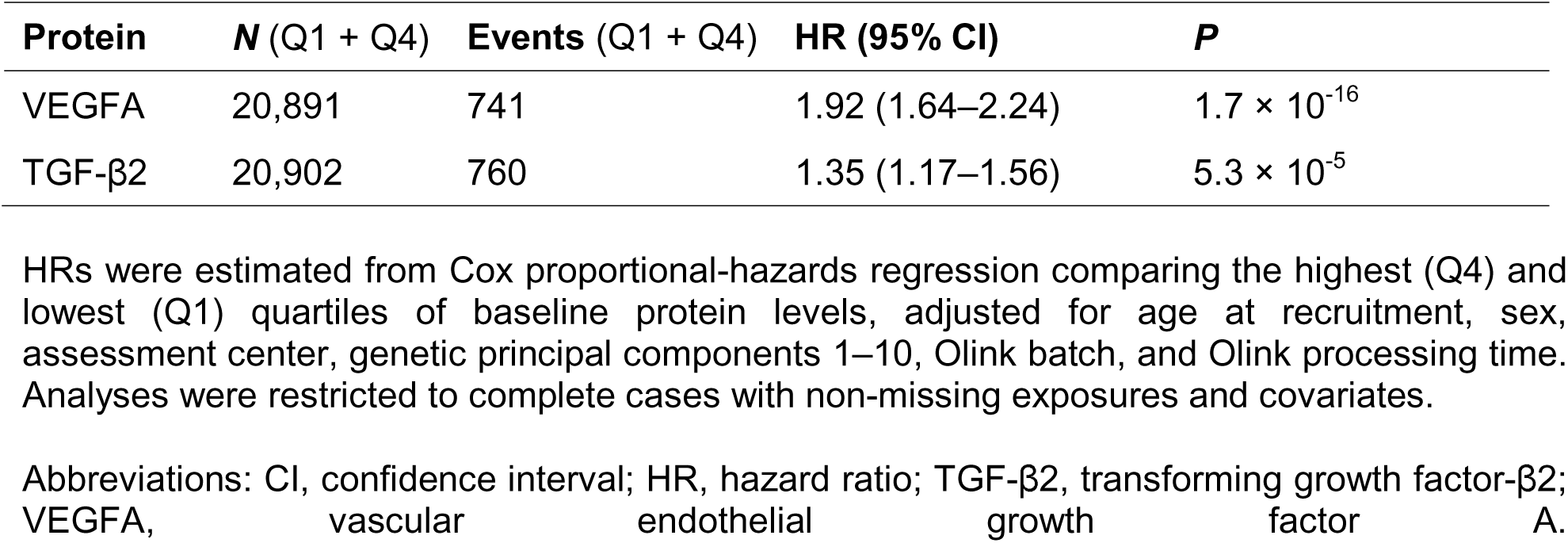
Associations of baseline VEGFA and TGF-β2 levels with incident type 2 diabetes in the UK Biobank (Q4 vs Q1 comparison).

### Druggability and therapeutic tractability

Given the roles of the genes *VEGFA* and *TGFB2* in adipose tissue biology and metabolic traits, we assessed their therapeutic potential for cardiometabolic diseases using the druggable genome^44^ and Open Targets Platform^45^. Both genes were classified as Tier 1 targets in the druggable genome, indicating that their protein products are modifiable by existing or investigational drugs. According to the Open Targets Platform, VEGFA had 11 drugs undergoing clinical trials, seven of which had reached phase 4 and five of those seven had completed phase 4.

## Discussion

In this study, we systematically mapped genetic variants to adipokine-coding genes and combined this information with multi-ancestry fine-mapping of 1,669 traits to pinpoint putatively causal variants. Clustering of these variants identified distinct IR signatures. Among them, a Thyroid-adiposity cluster was composed of *VEGFA* variants associated with adipose-related IR signals and thyroid function traits, and a Lipodystrophy cluster comprised variants in *TGFB2*, *VEGFA*, *VEGFB*, *NUCB2*, and *FGF21*, which were associated with lower body fat, abnormal glycemic traits, and high risk of metabolic diseases. Furthermore, regulatory annotations of variants within these two clusters provided supporting evidence consistent with each cluster’s mechanism. In complementary observational analyses in the UK Biobank, higher circulating VEGFA and TGF-β2 levels were associated with greater central adiposity, more adverse lipid profiles, and increased risk of incident T2D, phenocopying the patterns suggested by the genetic data. Finally, druggability assessment highlighted the adipokine-pathway genes, *VEGFA* and *TGFB2*, as having existing or potentially tractable therapeutic targets. Together, these findings demonstrate the existence of genetically driven, mechanistically distinct subtypes of IR, highlighting *VEGFA* and *TGFB2* as key players in two such pathways and emphasizing their potential translational relevance.

Previous efforts to classify the heterogeneity of IR and T2D through genetic clustering, such as those by Sevilla-González et al.^18^ and by Udler et al.^17^, grouped loci by shared trait associations and underlying biological pathways. Udler et al. identified a subgroup of IR-associated variants resembling monogenic familial partial lipodystrophy, characterized by high fasting insulin and TG but low HDL-C and reduced adiposity, which are features that parallel the Lipodystrophy cluster in our study. Variants within this cluster were associated with higher WHR, fasting insulin, and cardiometabolic risk, but lower BMI and BFP, supporting the concept that IR can occur independently of obesity and underlie “metabolically unhealthy lean” phenotypes. Sevilla-González et al. reported a fasting-insulin-lipodystrophy cluster near *PPARG*, *LYPLAL1, IRS1*, and *DNAH10*, genes previously implicated in lipodystrophy^49^. The recurrence of this lipodystrophy-like signal across multiple studies^24–27^ highlights a fundamental mechanism whereby impaired subcutaneous fat storage drives ectopic lipid accumulation and IR. Our findings reinforce this concept and extend it by implicating adipokine-related genes such as *TGFB2* and *VEGFA*, not classically recognized in lipodystrophy, as potential effector genes in this pathway.

TGF-β2 is a cytokine traditionally best known for development and immune roles, but recent data highlighted its metabolic functions. *TGFB2* mutations have been previously shown to lead to thoracic aortic aneurysm in mice^50^ and in humans^51^. In mice, adipose tissue secretes TGF-β2 in response to exercise, improving glucose tolerance^30^. These observations imply that higher TGF-β2 activity may benefit insulin sensitivity and suggest potential actionability for IR.

*VEGFA* plays a central role in angiogenesis and vascular homeostasis^52^. Overexpression in mice promoted the expansion of beige fat within subcutaneous white adipose tissue and conferred metabolic benefits during periods of fat mass gain^53,54^. *VEGFA* variants were also associated with higher WHRadjBMI^55^, aligning with our findings. In our study, *VEGFA* mapped to two distinct clusters, Lipodystrophy cluster and Thyroid-adiposity cluster, indicating a complex phenotype. Consistently, rs6905288 from the Lipodystrophy cluster was in strong LD (*r^2^* = 0.69) with rs998584, a top signal for fat mass ratio GWAS^56^ (a biomarker of lipodystrophy), and had prior links to HDL-C, TG, and WHRadjBMI^57,58^. By contrast, the thyroid–adipose cluster appears to represent a novel insight not evident in earlier clustering efforts.

The other genes found in the Lipodystrophy cluster included *VEGFB*, *NUCB2*, and *FGF21. VEGFB* enhances adipose tissue angiogenesis and glucose metabolism, improving insulin function and potential weight loss in obese mice^59^, and was proposed for treating IR and T2D in humans^60^. *NUCB2* is associated with hypertension in mice studies^61,62^, with concordant findings found in East Asian^63^ and European populations^64^. Furthermore, rs1073443 in *NUCB2* is highly associated with myeloperoxidase, a biomarker associated with atherosclerotic cardiovascular events^65^. Finally, *FGF21* has been linked to obesity^66^ in mice and, in variant clustering by Sevilla-González et al^18^, with lipid clusters.

Our study built on earlier clustering efforts which used only lead GWAS variants and assumed the nearest gene or a biologically obvious gene was responsible. In contrast, we empirically linked variants to genes using cS2G and validated cluster variants through orthogonal evidence, including tissue-specific eQTL data, chromatin accessibility profiles, and regulatory interaction maps, to identify promoter-interacting regions. This convergence of genetic mapping and epigenomics strengthened confidence that the clusters reflect true mechanisms, helping address the broader challenge described by Metz et al.^15^ of moving from variant to function, particularly in adipose tissue.

These advantages were evident in defining the Lipodystrophy cluster based on multi-ancestry fine-mapping results. Five variants mapped to *TGFB2* in this cluster may potentially act through long-range regulation or alternative splicing. Among them, rs4846565 showed the strongest negative association with BMI and BFP and the strongest positive association with T2D. This variant has been reported previously^31–33^, and our fine-mapping analysis likewise indicated it as the causal variant for the MVP GWAS weight, TG, baseline self-reported T2D, and diabetes mellitus supporting its relevance to glycemic control (**Supplementary Table 5**). Consistent with these findings, rs4846567 (chr1:219577375), a variant in high LD with rs4846565 in Europeans (*r^2^* = 0.74), has been associated with increased weight loss, reduced hunger^67^, and abdominal obesity in East Asians^68^. Notably, these five *TGFB2* variants are located closest to *LYPLAL1*, which has also been linked to obesity^55^. rs4846565 (chr1:219548762-A) was significantly associated with altered splicing of the antisense RNA *LYPLAL1-AS1* in subcutaneous adipose tissue, with the alternative allele reducing the intron excision rate of chr1:219449233:219458858 (normalized effect size = -0.45, *P* = 3.3 × 10^-15^). Further investigations are required to elucidate *trans*-regulatory interactions, resolve distal effects, and characterize possible tissue- or context specificity at this locus.

Prior variant clustering studies did not report a thyroid-linked IR cluster, possibly due to differences in the loci or traits considered. This underscores the value of our analysis in uncovering an additional layer of IR heterogeneity beyond previously known clusters. The Thyroid-adiposity cluster points to an IR mechanism involving endocrine crosstalk between adipose tissue and the thyroid axis. *VEGFA* variants, for instance, were associated with increased WHRadjBMI and TSH levels but showed protective effects against T2D. Although elevated WHRadjBMI and TSH usually indicate higher T2D risk via central obesity and hypothyroidism^69,70^, these variants deviated from that pattern. This suggests a unique biological pathway whereby higher TSH or altered adiposity distribution confers protection against diabetes, possibly through indirect or compensatory pathways. A *VEGFA*-proximal variant may alter adipose secretion of VEGFA or related adipokines, affecting systemic metabolism and thyroid hormone signaling. While the precise link between *VEGFA* and thyroid function remains unclear, our findings propose a thyroid–adipose axis contributing to IR, broadening IR genetics beyond classical lipodystrophic pathways.

From a translational perspective, recognizing these clusters may help prioritize more mechanism-informed therapeutic strategies. For example, given their adipose-specific expression signatures, individuals whose genetics indicate a lipodystrophy-like IR mechanism might benefit from therapies that improve adipose function or mimic adipokine replacement, similar to how metreleptin therapy markedly improved insulin sensitivity and hypertriglyceridemia in lipodystrophy patients^71^. Consistent with this translational potential, several VEGFA-targeting drugs (e.g., aflibercept and bevacizumab) are FDA-approved for retinal disorders such as diabetic retinopathy and diabetic macular edema^46,47^, and may be repurposed given VEGFA’s role in adipose angiogenesis and insulin sensitivity. TGF-β2 has four drugs targeting it undergoing clinical trials, with one drug, luspatercept, already having reached phase 4. While TGF-β2 lacks approved therapies, inhibitors of the TGF-β pathway are being developed for blood-related disorders including β-thalassemia^48^. However, the present findings should not be taken to imply that existing VEGFA-targeting therapies are directly repurposable for metabolic disease. *VEGFA* biology is highly tissue- and context-dependent, and systemic inhibition may be inappropriate given known on-target vascular effects. Instead, our results nominate *VEGFA*- and *TGFB2*-related pathways as candidates for mechanistic follow-up and for future exploration of tissue-selective or pathway-selective modulation.

This study has limitations. First, while multi-ancestry fine-mapping improved resolution, reliance on external LD panels may introduce artifacts when reference LD structure diverged from that of the GWAS samples^72^. Nevertheless, we used s-values^73,74^ to assess LD consistency and results suggested low LD mismatch between summary statistics and reference panels (**Supplementary Note 2**). Second, while the MVP cohort provided one of the largest publicly available GWAS datasets for individuals of African ancestry, the East Asian sample size was limited, and greater diversity is needed. Third, although the cS2G strategy increased confidence in V2G assignment, interpreting the function of variants in non-coding regions remains challenging. For instance, cS2G and Open Targets V2G do not treat non-coding elements, such as lincRNA, as targets. Moreover, as an *in-silico* method, cS2G is inherently probabilistic and does not prove causality. Therefore, functional experiments remain essential to validate the causal gene and mechanism at each locus.

In summary, systematic V2G mapping and multi-ancestry fine-mapping of adipokine genes, coupled with variant clustering, revealed that genetic predisposition to IR could be classified into distinct mechanistic clusters rooted in specific biological pathways. We identified *TGFB2* as a key regulator in non-obese, lipodystrophy-like IR. We also showed potential diverging mechanisms through which *VEGFA* exerted its regulatory effects. These insights expanded understanding of how adipose dysfunction contributes to metabolic disease and informed precision medicine approaches, including prioritizing *VEGFA*- and *TGFB2*-related pathways for mechanism-guided translational follow-up and future therapeutic development tailored to genetically defined IR subtypes.

## Methods

### GWAS outcomes from the Million Veteran Program (MVP)

The MVP cohort provides up to 2,068 traits across multiple ancestries. Details on genotyping, imputation, and quality control can be found in the original study^19^. See **Supplementary Note 1** for a description of the GWAS.

### Linking SNPs to genes using cS2G

We leveraged the combined SNP-to-gene (cS2G) strategy^16^ which linearly combines seven constituent S2G strategies together to link variants to genes. cS2G maximized recall while ensuring that precision remained at or above 0.75. The seven S2G methods ordered based on weight from highest to lowest constitute the following: exon, promoter, eQTLGen blood fine-mapped cis-eQTL, GTEx fine-mapped cis-eQTL, EpiMap enhancer–gene linking, ABC, and Cicero blood/basal. More details on cS2G can be found in the original study^16^. In total, 3,510,384 variants had a cS2G annotation score.

### Defining loci for multi-ancestry fine-mapping

We used European, African, and East Asian ancestry loci defined in Supplementary Table 3 of the original MVP study^19^ but redefined them for multi-ancestry fine-mapping. For loci with only a single lead variant and no other variants in the region, we padded ±250kb on either side of the lead variant to redefine the region for fine mapping. To curate loci for multi-ancestry fine-mapping, we combined loci from all three ancestries by merging any overlapping regions into a single locus. In total, we identified a total of 753 traits across all three ancestries which had at least one locus encompassing 45,974 non-overlapping regions (33,179 loci shared between all three ancestries and 12,795 loci shared between European and African ancestries).

### Cross-population fine-mapping with SuSiEx

SuSiEx^20^ is a multi-ancestry fine-mapping method which can combine population-specific GWAS from different ancestries and along with their specific LD reference panels to improve power and enhance fine-mapping accuracy. We used summary level GWAS from MVP and 1000 Genomes Project phase 3 samples^75^ LD reference panels for European, African, and East Asian ancestries as input. Analyses were performed using SuSiEx-v1.1.2 with default settings (MAF_THRESHOLD=0.005, --n_sig=5) but we set --keep-ambig=True and --mult-step=True which performs a multi-step model fitting approach. We used 95% as the coverage level of credible sets.

To assign a representative gene to each credible set, we aggregated variant-level posterior inclusion probabilities (PIPs) across all variants in the credible set for each gene linked by cS2G. Specifically, for gene *g* in credible set *C*, we computed a gene-level score as:

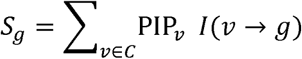

where *PIP_v_* is the posterior inclusion probability of variant *v*, and *I* (*v* → *g*) is an indicator equal to 1 if variant *v* was linked to gene *g* by cS2G and 0 otherwise. Thus, if a variant was linked to multiple genes, its full PIP contributed to each linked gene. The representative effector gene for the credible set was then defined as the gene with the largest summed PIP, *g*\* = arg max*_g_ S_g_*.

SuSiEx uses the POST-HOC_PROB_POP output to assign a Post hoc probability (that a credible set manifests as causal in a population) for each ancestry to the highest PIP variant in each credible set. If a credible set had a post hoc prob > 0.8 in two or more ancestries, we considered multi-ancestry fine-mapping to have been performed. Credible sets with a post hoc probability > 0.8 in only one ancestry were considered to have been fine-mapped in that ancestry only.

### Quantifying LD mismatch

We used the estimate_s_rss() function from the susieR^73,74^ package to estimate the s value which quantifies the amount of LD mismatch. The s value ranges between 0 (no LD mismatch) to 1 (complete LD mismatch) with larger values indicating stronger inconsistency between GWAS z scores and the LD matrix. If a loci had an s value > 0.1 we considered this as evidence of LD mismatch.

### Adipokines

Defining a comprehensive list of adipokines is complex due to disparate criteria for inclusion. Nevertheless, we compiled a list by considering factors such as predominant expression in adipose tissue, evidence of secretion, and functional relevance using multiple sources^2,76,77^ and literature searches (**Supplementary Table 4**).

We created network plots by systematically extracting gene-trait pairs from adipokine coding genes and excluding pairs whose traits included (1) those related to height, blood cells, and family history; (2) biomarker traits which pertain to maximum/minimum values for each individual across all visits (only the Mean value trait was assessed). This resulted in 228 pairs of 40 adipokine genes and 107 traits.

### Clustering of genetic variants

To cluster genetic variants mapped to adipokine genes, we performed the following steps (**Supplementary Figure 2**).

1. *Select disease-relevant adipokine genes: the list of gene-trait pairs is used to select a list of adipokine genes that are relevant for diseases.* The 228 adipokine-trait associations from 40 adipokines and 107 traits included a unique list of 163 SNPs. These SNPs were the top-PIP-scoring variants for adipokine gene-trait pairs. Of these, 156 SNPs were present in the waist-to-hip ratio adjusted for BMI (WHRadjBMI) European ancestry GWAS from Pulit et al.57 and were thus analyzed.
2. *Characterization: create a matrix where rows are the top PIP variant (for these adipokine genes), and columns are cardiometabolic-related continuous and binary traits.* To assess the direction of SNPs, we included nine cardiometabolic traits and aligned all effect alleles to increase WHRadjBMI.

- The six continuous traits were body mass index (BMI), body fat percentage (BFP), ratio of visceral adipose tissue (VAT) to gluteofemoral adipose tissue (GFAT) (VATtoGFAT), HbA1C, fasting glucose (FG), and fasting insulin (FI).
- The three binary traits were stroke (any ischemic stroke), coronary artery disease (CAD), and type 2 diabetes (T2D).
3. *Clustering of 156 genetic variants* Since the nine cardiometabolic-related traits were mostly curated from external sources, not all 156 SNPs were present in some traits. To address this, we set missing values to 0 then used the NbClust() package in R (https://cran.r-project.org/web/packages/NbClust/index.html) to perform hierarchical clustering to select the optimal number of clusters. Next, we performed imputation of those missing values using the ClustImpute() R package (https://cran.r-project.org/package=ClustImpute) as done previously^78^ while using the number of clusters from NbClust() as the input number of clusters. We then performed NbClust() again on the outputted imputed results to obtain the optimal number of clusters for visualization. We used the R package pheatmap for the heatmap visualization.

#### Regulatory role of lipodystrophy-like insulin resistance genetic variants linked to TGFB2

To assess the regulatory potential of *TGFB2* variants associated with a lipodystrophy-like phenotype, we integrated epigenomic and chromatin interaction datasets relevant to adipose tissue. Chromatin accessibility was evaluated using ATAC-seq data from the ENCODE^34^ consortium in subcutaneous and visceral adipose tissues with regulatory annotations obtained from RegulomeDB^35^. Active histone modifications, including H3K27ac and H3K4me3, were examined to infer enhancer and promoter activity.

To determine physical interactions between distal regulatory elements and gene promoters, promoter Capture Hi-C (pCHi-C) data from human preadipocytes were analyzed, as previously described by Garske et al.^37^ These data were used to identify long-range chromatin looping events between variant-containing loci and the *TGFB2* promoter. To do so, we padded 30kb on either side of the rs61823192 variant and assessed for overlap with a 1kb upstream and 100bp downstream region of the *TGFB2* promoter. We used the plotgardener R package to create plots.

### Regulatory evidence for *VEGFA*

#### Expression and splicing quantitative trait loci

To investigate the regulatory effects of four *VEGFA*-associated variants on gene expression and alternative splicing across tissues, we utilized publicly available transcriptomic datasets. Expression quantitative trait loci (eQTL) and splicing QTL (sQTL) analyses were conducted using the Genotype-Tissue Expression (GTEx) Project v8 database^38^, which provides multi-tissue transcriptomic data. We queried the variants across all available tissues, focusing on their associations with expression levels and splicing patterns of *VEGFA*. Statistical significance was determined based on GTEx-provided *P* values.

Additionally, to assess adipose tissue-specific regulatory effects and validate variant-gene expression associations in a metabolically relevant context, we examined variant associations in the AdipoExpress eQTL meta-analysis by Brotman et al.^39^, comprising 2,344 subcutaneous adipose tissue samples. AdipoExpress provides publicly available full marginal eQTL summary statistics in European ancestry with analyses performed within 1Mb of the canonical gene transcription start site. We queried variants in AdipoExpress and determined significant associations for the gene of interest as associations with *P* < 0.05.

#### Epigenomic evidence

To evaluate the potential regulatory functions of the identified *VEGFA* intronic variants, we queried each variant in RegulomeDB^35^, a database that integrates diverse epigenomic and transcriptional datasets to predict regulatory potential. RegulomeDB assigns ranks based on the presence of functional annotations such as transcription factor (TF) binding, DNase hypersensitivity, chromatin accessibility, eQTLs, and known regulatory motifs. Variants were assigned both a categorical rank (1a–7) and a numerical score (0–1), with lower ranks and higher scores indicating stronger regulatory evidence. These annotations were used to prioritize variants with the highest likelihood of regulatory function.

### Phenome-wide association study analyses

Phenome-wide association studies (PheWAS) were conducted to evaluate the clinical relevance of four *VEGFA* intronic variants across a wide range of human traits. Variant-trait associations were queried using the Common Metabolic Diseases Knowledge Portal (https://hugeamp.org/), which aggregates large-scale GWAS summary statistics for metabolic traits. Associations were assessed across hundreds of phenotypes, with particular attention to thyroid-related traits such as thyroid-stimulating hormone (TSH) (effective sample size, *n* = 635,908) and free thyroxine (FT4) levels (*n* = 117,166). Effect sizes (β) and *P* values were extracted to identify the most significant associations.

#### Dissecting the regulatory complexity at the VEGFA locus reveals evidence of functional pleiotropy

To investigate the regulatory landscape of the *VEGFA* locus and delineate the functional distinctions between *VEGFA* variants grouped into Cluster 2 (Thyroid-adiposity cluster) and Cluster 3 (Lipodystrophy cluster), we conducted a series of integrative genomic analyses. Specifically, we assessed their tissue-specific regulatory effects and potential functional pleiotropy. eQTL analyses were performed using publicly available data from GTEx v8 and AdipoExpress as described in earlier sections of the methods. For each variant, we assessed the presence of *cis*-eQTL associations with *VEGFA* and other proximal genes across multiple tissues, with particular emphasis on subcutaneous adipose and thyroid tissues. Variants were considered eQTLs if they exhibited a nominal association with gene expression (*P* < 0.05). Colocalization analyses were extracted from the CoLocus browser (https://amp.colocus.app/) integrated into the Common Metabolic Diseases Knowledge Portal (https://hugeamp.org/) for *VEGFA.* Colocalization assesses the probability of shared causal variants between GWAS traits and gene expression signals (here AdipoExpress eQTL). A threshold of ≥ 0.7 was used to suggest strong colocalization evidence.

Regional association plots and tissue-specific eQTL landscapes were examined using locuszoom to identify distinct regulatory peaks and their corresponding lead variants. Chromosomal positions and allelic configurations were derived from the GRCh38 reference genome. Lead *cis*-eQTLs for *VEGFA*, *LINC02537*, and *LINC01512* were identified in both thyroid and adipose tissues, and their expression profiles were compared across clusters. Phenome-wide association analyses (PheWAS) were performed to evaluate potential trait associations with Cluster 3 variants in thyroid-related phenotypes.

### Observational analyses in the UK Biobank

#### Cross-sectional associations between circulating protein levels and adiposity-related traits

Using data from the UK Biobank cohort, we performed multivariable linear regression to evaluate the associations between baseline circulating levels of VEGFA and TGF-β2 and adiposity-related traits, including BMI (UK Biobank data field: 21001), WHR (data field: waist circumference, 48; hip circumference, 49), and the triglyceride (TG) to high-density lipoprotein cholesterol (HDL-C) ratio (data field: TG, 30870; HDL-C, 30760). Protein concentrations were quantified using the Olink Explore 3072 assay (data field: 30900). Both the protein (exposure) and each trait (outcome) were standardized to mean 0 and SD 1 so that β represents the SD change in the trait per 1-SD higher protein level. Analyses were adjusted for age at recruitment (data field: 21022), sex (data field: 31), assessment center (data field: 54), genetic principal components (PCs) 1–10 (data field: 22009), Olink measurement batch (resource: 1016), and Olink processing time (resource: 1016). When WHR was the outcome, BMI was added as an additional covariate to account for overall adiposity. All analyses were based on complete cases. For multiple testing across the six linear-regression tests (two proteins × three traits), a Bonferroni-corrected threshold of *P* < 0.0083 (0.05/6) was considered statistically significant, with *P* < 0.05 considered nominally significant.

#### Associations between circulating proteins and incident type 2 diabetes

We examined whether baseline VEGFA and TGF-β2 levels were associated with the incidence of type 2 diabetes (T2D) using multivariable Cox proportional-hazards regression implemented in the coxph() function (survival v3.5.5 R package), using longitudinal data from the UK Biobank.

For each protein, participants were categorized into quartiles according to baseline concentrations, and hazard ratios were estimated comparing the highest (Q4) with the lowest (Q1) quartile. T2D events were identified from hospital inpatient records using ICD-10 code E11 including all subcategories; admission dates were preferentially taken from the recorded admission date and otherwise from the episode start date. Participants with a T2D diagnosis on or before the baseline assessment date (data field: 53) were excluded. Follow-up time was calculated from baseline to the earliest of T2D diagnosis, loss to follow-up (data field: 191), death (derived from the national death registry), or 31 May 2022. The Cox regression used the same covariate set as the cross-sectional analyses (age, sex, assessment center, genetic PCs 1–10, and Olink processing variables) and was restricted to complete cases with non-missing exposures and covariates. A Bonferroni-corrected threshold of *P* < 0.025 (0.05/2) was considered statistically significant.

The UK Biobank study was approved by the North West Multicentre Research Ethics Committee, and all participants provided written informed consent. Analyses were conducted under UK Biobank application 73958.

### Druggability assessment of replicated putatively causal protein-phenotype pairs

To explore therapeutic potential, we investigated drug repurposing opportunities for *TGFB2* and *VEGFA*. Given their established roles in vascular biology and tissue remodeling, we assessed whether existing pharmacological agents targeting these pathways could be repositioned to modulate insulin resistance or related phenotypes since this may allow translation of genetic findings into potential therapies. We used the druggable genome from Finan et al.^44^ and Open Targets Platform^45^ to query these protein-coding genes following established methodology from our earlier study^79^. Briefly, Finan et al. classified human protein-coding genes into three tiers based on their druggability to support target discovery and repurposing where Tier 1 included genes targeted by approved drugs or those in clinical development.

## Ethics declarations

All contributing cohorts obtained ethical approval from their institutional ethics review boards.

## Data availability

Million Veteran Program GWAS are available from the original study^19^

cS2G^16^: https://zenodo.org/records/10117202

AdipoExpress^39^: https://zenodo.org/records/13845120

RegulomeDB database^35^: https://www.regulomedb.org/regulome-search

PCHi-C data from Garske et al.^37^: https://www.ncbi.nlm.nih.gov/geo/query/acc.cgi?acc=GSE183770

Common Metabolic Diseases Knowledge Portal (https://hugeamp.org/)

## Code availability

We used R v4.4.1 (https://www.r-project.org/),

PLINK v1.9^80^ (http://pngu.mgh.harvard.edu/purcell/plink/),

Cytoscape (https://cytoscape.org/),

LocusZoom^81^ (https://my.locuszoom.org/),

SuSiEx v1.1.2^20^ (https://github.com/getian107/SuSiEx),

Plotgardener^82^ (https://phanstiellab.github.io/plotgardener/index.html)

## Acknowledgments

M.H. is supported by the Japan Student Services Organization (Graduate Scholarship for Degree-Seeking Study Abroad) and by the Watanabe Foundation (6th Toshizo Watanabe International Scholarship). T.L. is supported by start-up funding from the Office of the Vice Chancellor for Research and Graduate Education, School of Medicine and Public Health, and Department of Population Health Sciences at the University of Wisconsin-Madison. S.Y. is supported by the Japan Society for the Promotion of Science. The funders had no role in the study design, data collection and analysis, decision to publish, or preparation of the manuscript. We acknowledge Servier Medical Art (https://smart.servier.com/) for providing images that were used to create diagrams in this study.

## Author contributions

Conception and design: C.-Y.S., T.L., S.Y.

Methodology: C.-Y.S., M.H., T.L., S.Y.

Data curation: C.-Y.S.

Data Analysis: C.-Y.S., M.H.

Visualization: C.-Y.S., M.H., A.v.d.G.

Writing—Original Draft: C.-Y.S.

Writing—Review and Editing: all authors

Supervision: T.L., S.Y.

## Supplementary Figure Legands

**Supplementary Figure 1.**
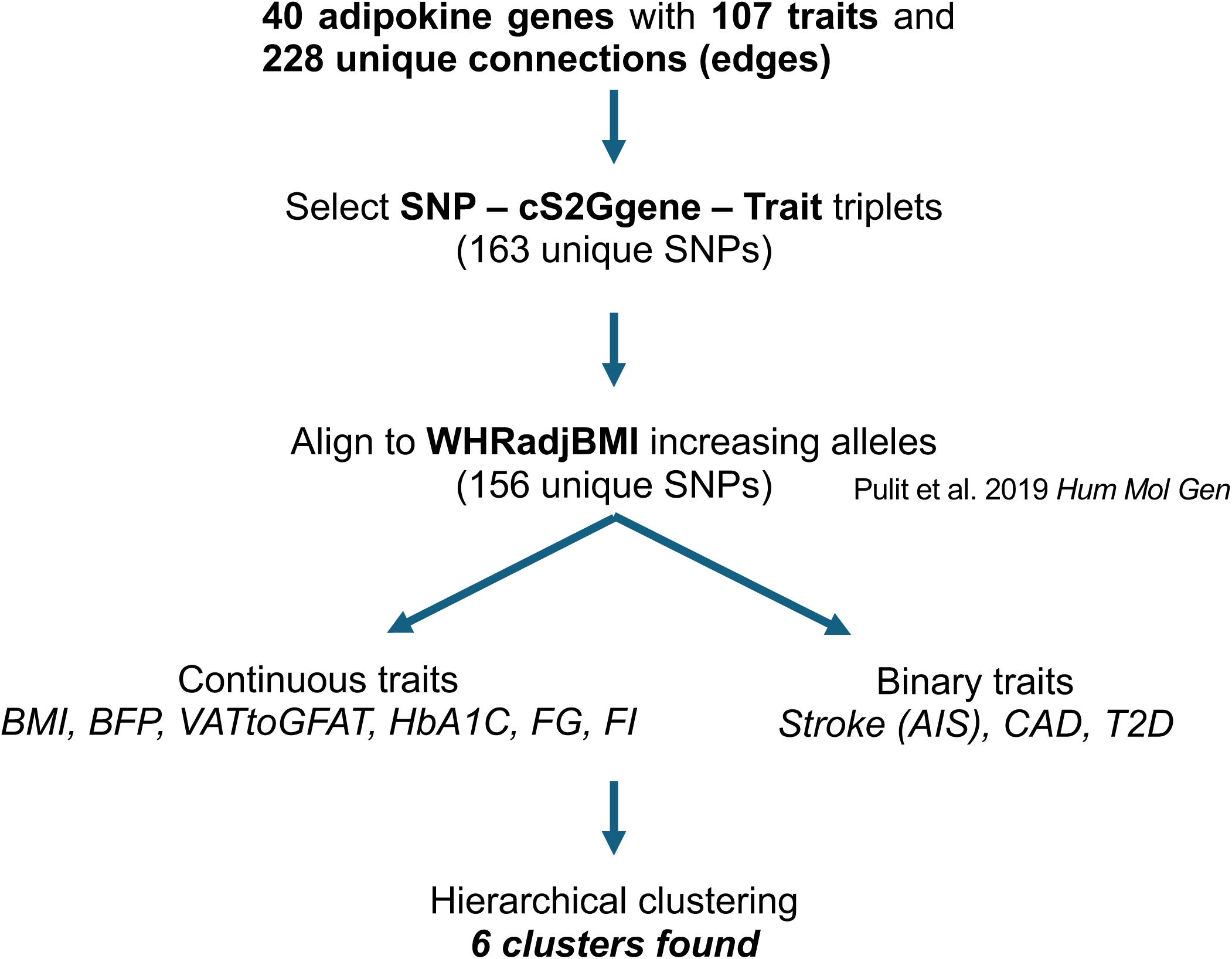
Flow diagram showing clustering of genetic variants of adipokine genes. Flow diagram summarizing the steps used to derive adipokine variant clusters. We started from 40 adipokine-encoding genes connected to 107 traits with 228 unique gene–trait edges. From these, we selected SNP–cS2G gene–trait triplets (163 unique SNPs) obtained from the variant-to-gene mapping pipeline and aligned all effect estimates to the waist-hip ratio adjusted for BMI (WHRadjBMI)–increasing allele using summary statistics from Pulit et al.^57^, resulting in 156 unique SNPs. Continuous traits included BMI, body fat percentage (BFP), visceral-to-gluteofemoral fat ratio (VATtoGFAT), glycated hemoglobin (HbA1c), fasting glucose (FG), and fasting insulin (FI), whereas binary traits included ischemic stroke (AIS), coronary artery disease (CAD), and type 2 diabetes (T2D). Hierarchical clustering of standardized SNP–trait associations across these nine cardiometabolic traits identified six distinct variant clusters.

**Supplementary Figure 2.**
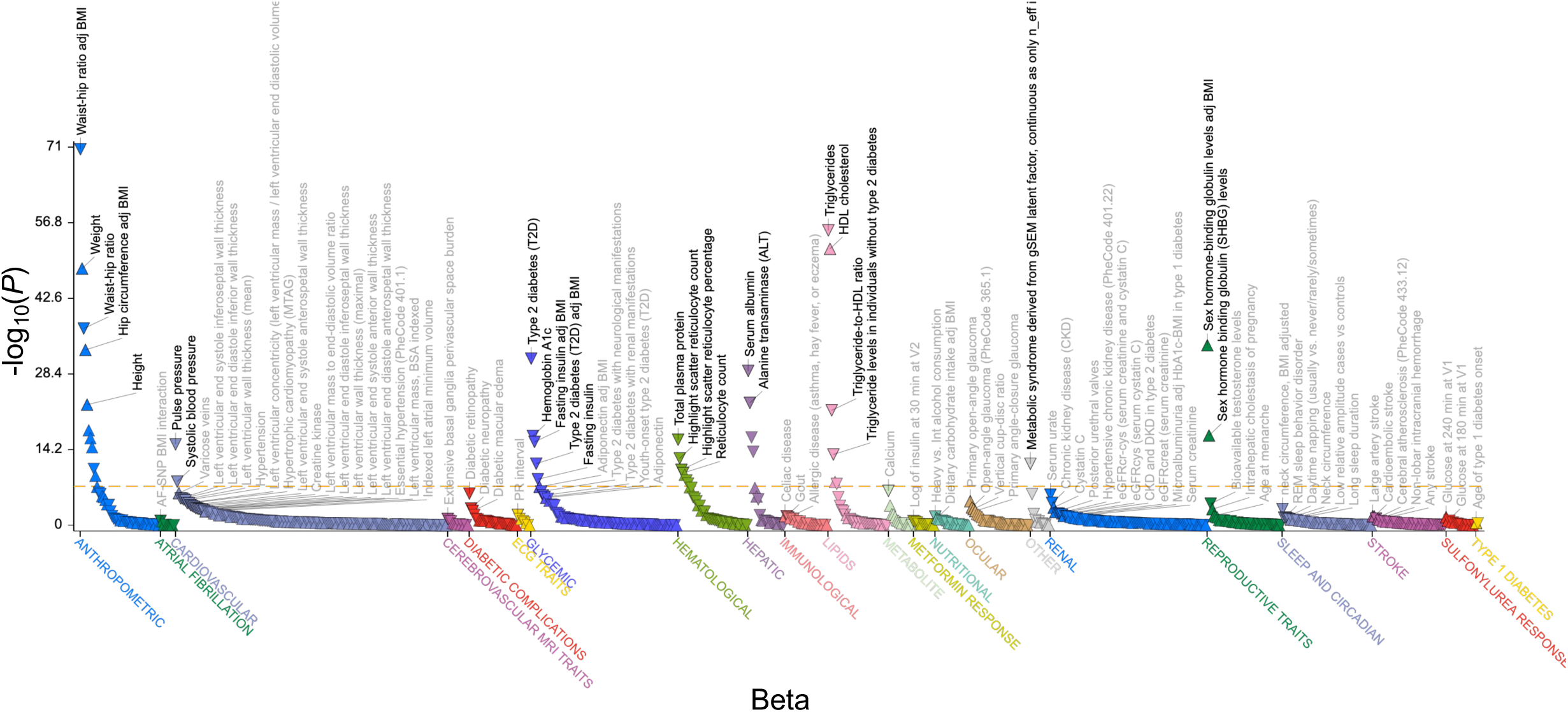
Phenome-wide association plot for rs4846565. Phenome-wide association results of rs4846565 (chr1:219,548,762 G>A) using data from the Common Metabolic Disease Knowledge Portal (https://hugeamp.org). Each point represents a single phenotype, with the x-axis showing the effect size (β) and the y-axis showing the corresponding -log10(*P*) value for SNP–trait associations across the phenome.

**Supplementary Figure 3.**
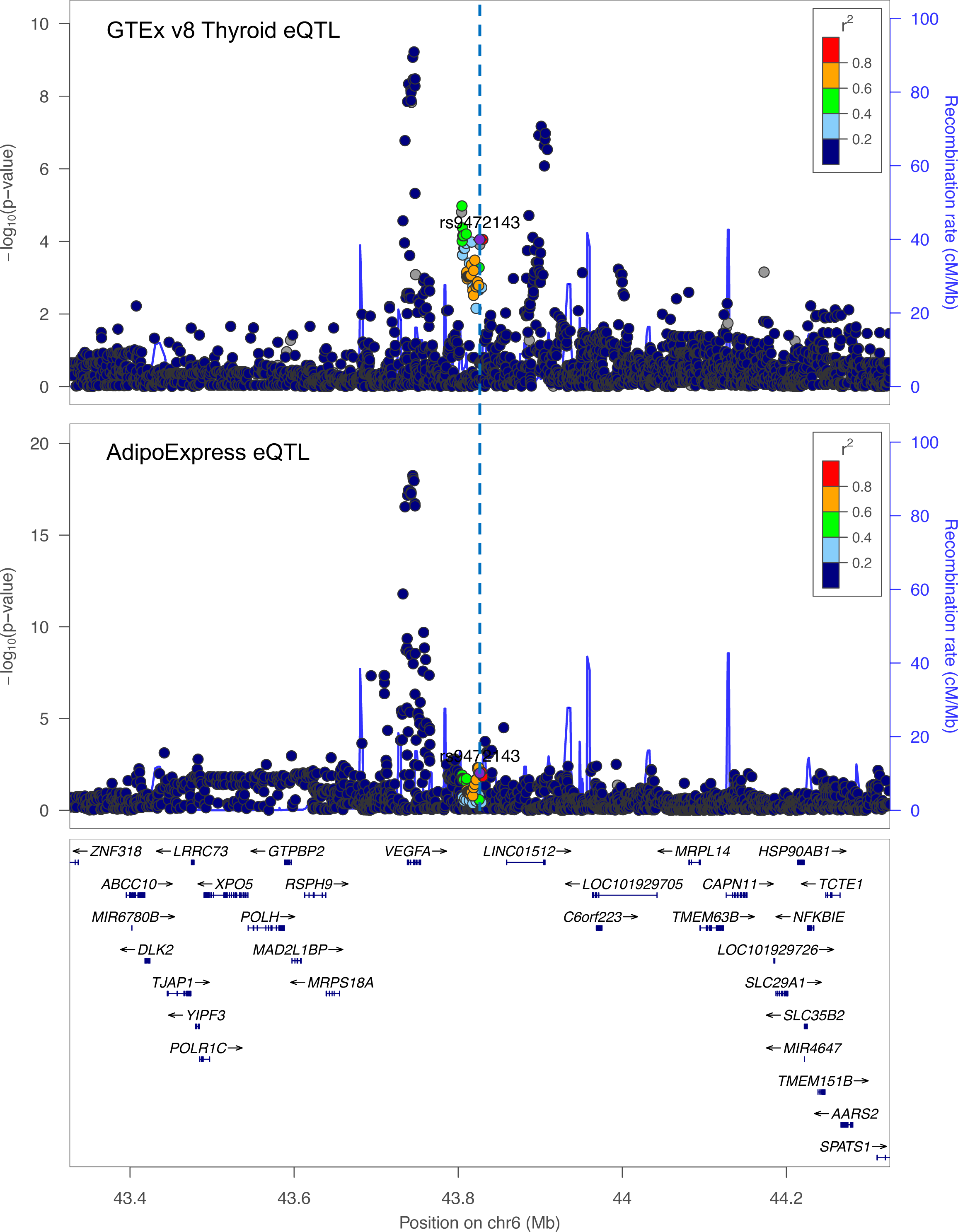
Regional *cis*-eQTL plot for rs9472143 in thyroid and adipose tissue. Regional association plots around rs9472143 at the *VEGFA* locus showing *cis*-eQTL signals in (top panel) thyroid tissue from GTEx v8 and (bottom panel) adipose tissue from the AdipoExpress study. For each variant in the region, the y-axis shows -log10(*P*) for association with *VEGFA* expression, and the x-axis shows chromosomal position on chromosome 6. Recombination rate (cM/Mb) is overlaid to indicate local linkage disequilibrium structure, and nearby genes are displayed at the bottom to provide genomic context around *VEGFA* and neighboring loci.

**Supplementary Figure 4.**
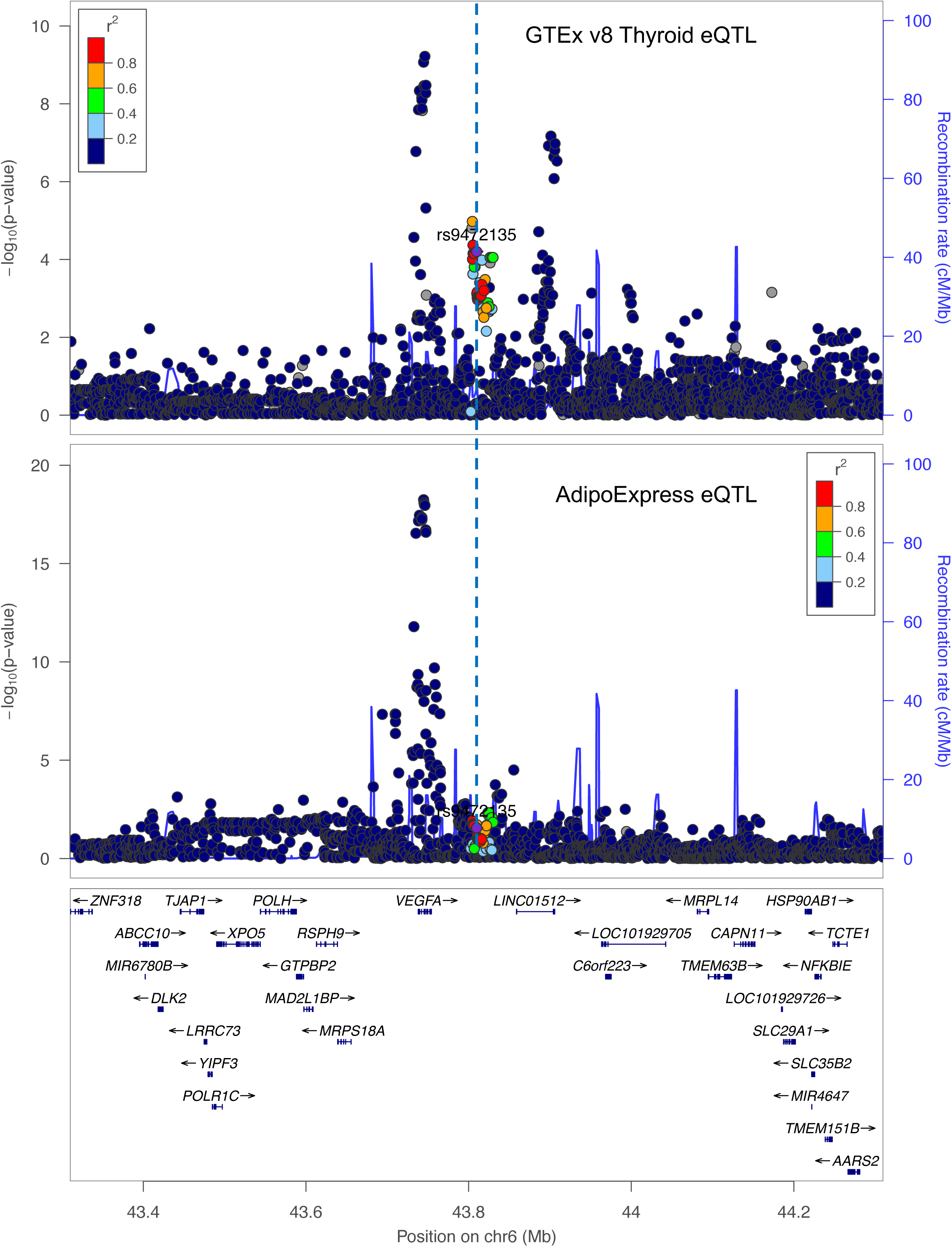
Regional *cis*-eQTL plot for rs9472135 in thyroid and adipose tissue. Regional *cis*-eQTL plots for rs9472135 at the *VEGFA* locus. The top panel shows GTEx v8 thyroid eQTL signals for *VEGFA*, and the bottom panel shows AdipoExpress adipose eQTL signals. Points represent variants in the region, with -log10(*P*) for association with *VEGFA* expression plotted against chromosomal position on chromosome 6. The overlaid recombination rate curve illustrates local LD patterns, and the gene track at the bottom marks *VEGFA* and neighboring genes, providing evidence for shared regulatory architecture across thyroid and adipose tissues.

**Supplementary Figure 5.**
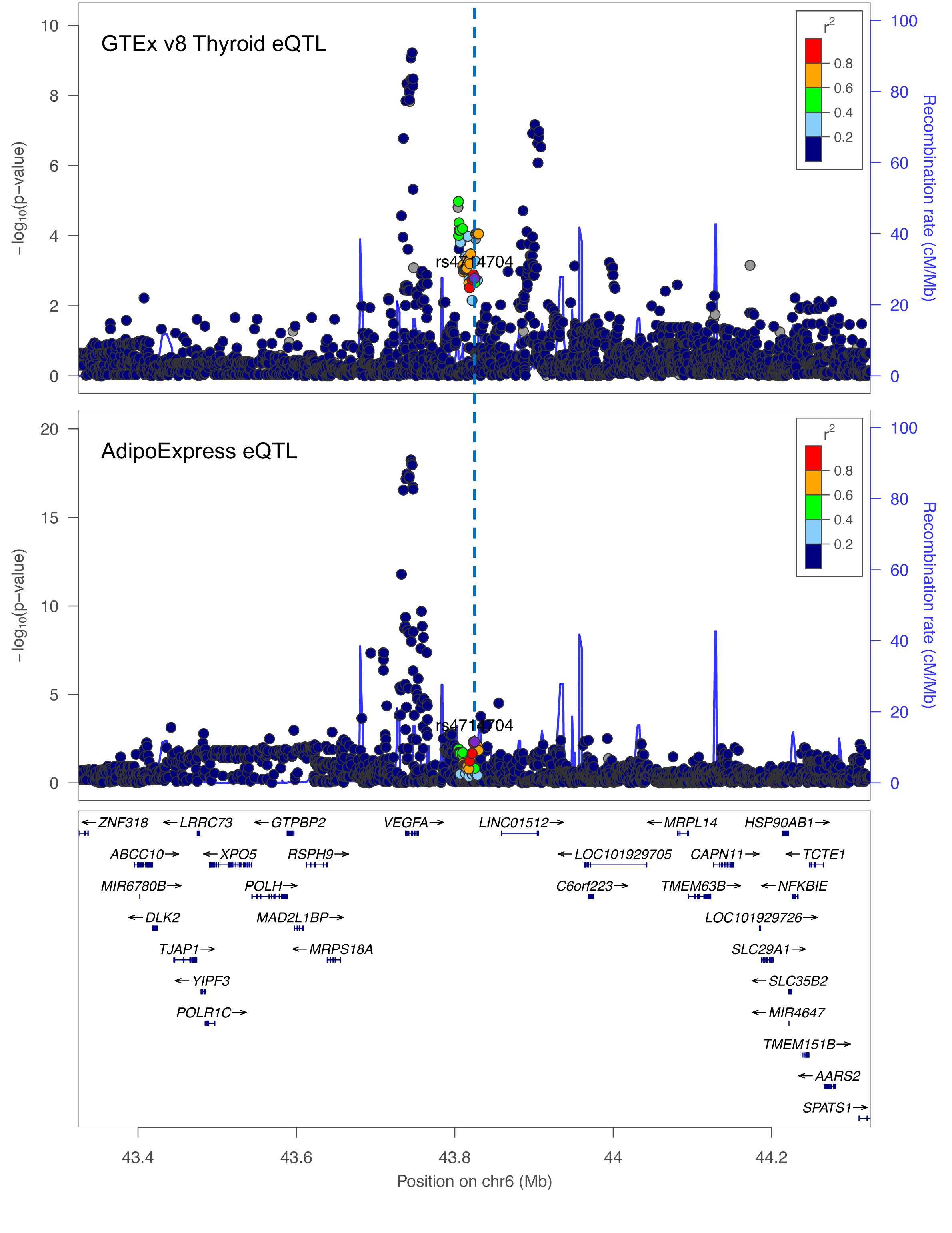
Regional *cis*-eQTL plot for rs4714704 in thyroid and adipose tissue. Regional eQTL association plots centered on rs4714704 at the *VEGFA* locus, showing *VEGFA cis*-eQTL signals in GTEx v8 thyroid (top) and AdipoExpress adipose tissue (bottom). Variants are plotted by chromosomal position (x-axis) and -log10(*P*) value for association with *VEGFA* expression (y-axis), with recombination rate (cM/Mb) superimposed.

**Supplementary Figure 6.**
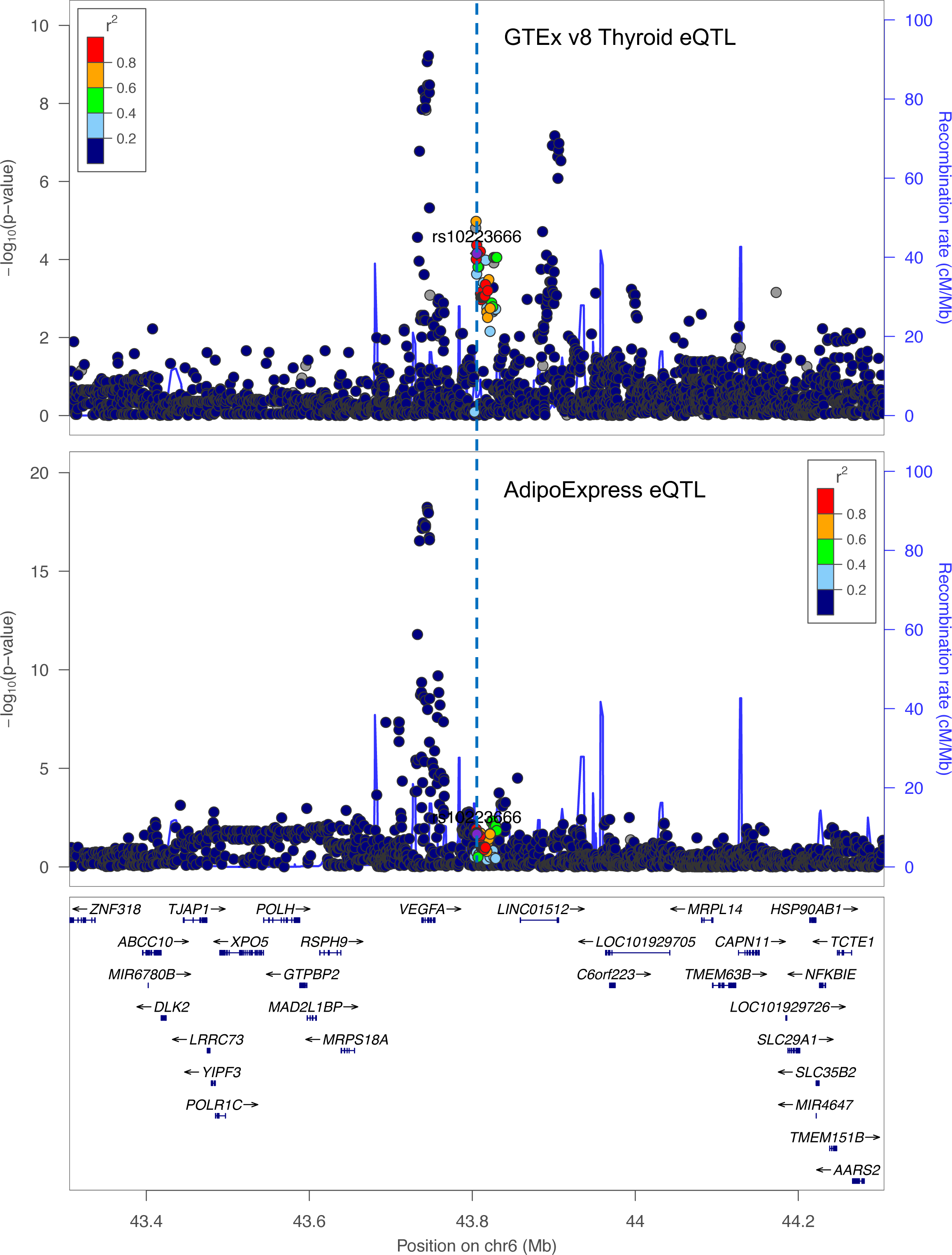
Regional *cis*-eQTL plot for rs10223666 in thyroid and adipose tissue. Regional *cis*-eQTL plots for rs10223666 at the *VEGFA* locus. GTEx v8 thyroid eQTL results are shown in the top panel, and AdipoExpress adipose eQTL results in the bottom panel. Each point represents a variant in the locus, with -log10(*P*) for association with *VEGFA* expression on the y-axis and chromosomal position on the x-axis. Recombination rate is displayed to illustrate LD patterns, and nearby genes are indicated at the bottom.

**Supplementary Figure 7.**
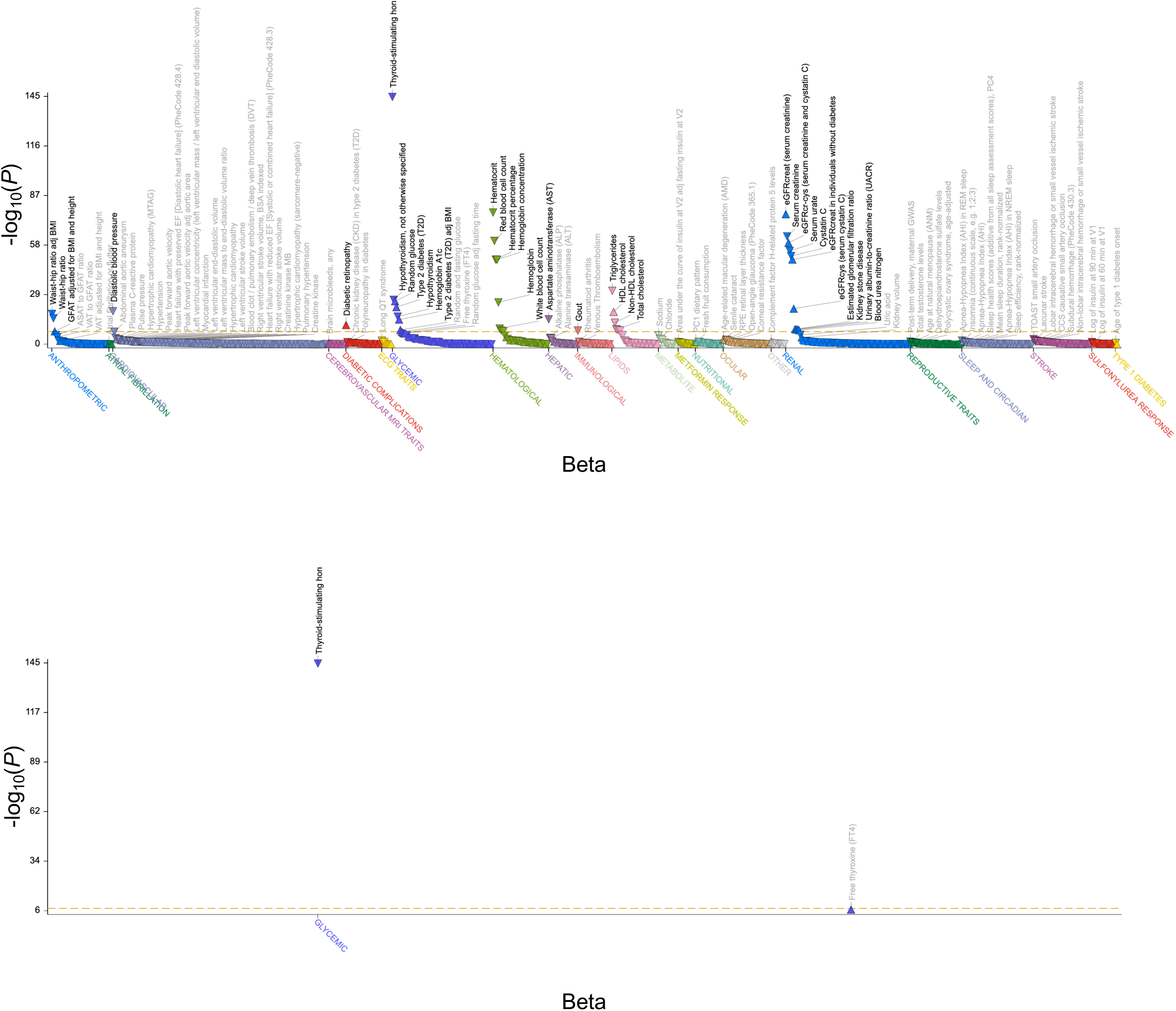
Phenome-wide association plot for rs9472143. Phenome-wide association results of rs9472143 using data from the Common Metabolic Disease Knowledge Portal (https://hugeamp.org) is shown on the top panel. The bottom panel shows a focused view displaying only the associations of rs9472143 with thyroid-stimulating hormone (TSH) and free thyroxine (FT4), plotted on the same scales, highlighting the specific effects of this variant on thyroid function traits. Each point represents a single phenotype, with the x-axis showing the effect size (β) and the y-axis showing the corresponding -log10(*P*) value for SNP–trait associations across the phenome.

**Supplementary Figure 8.**
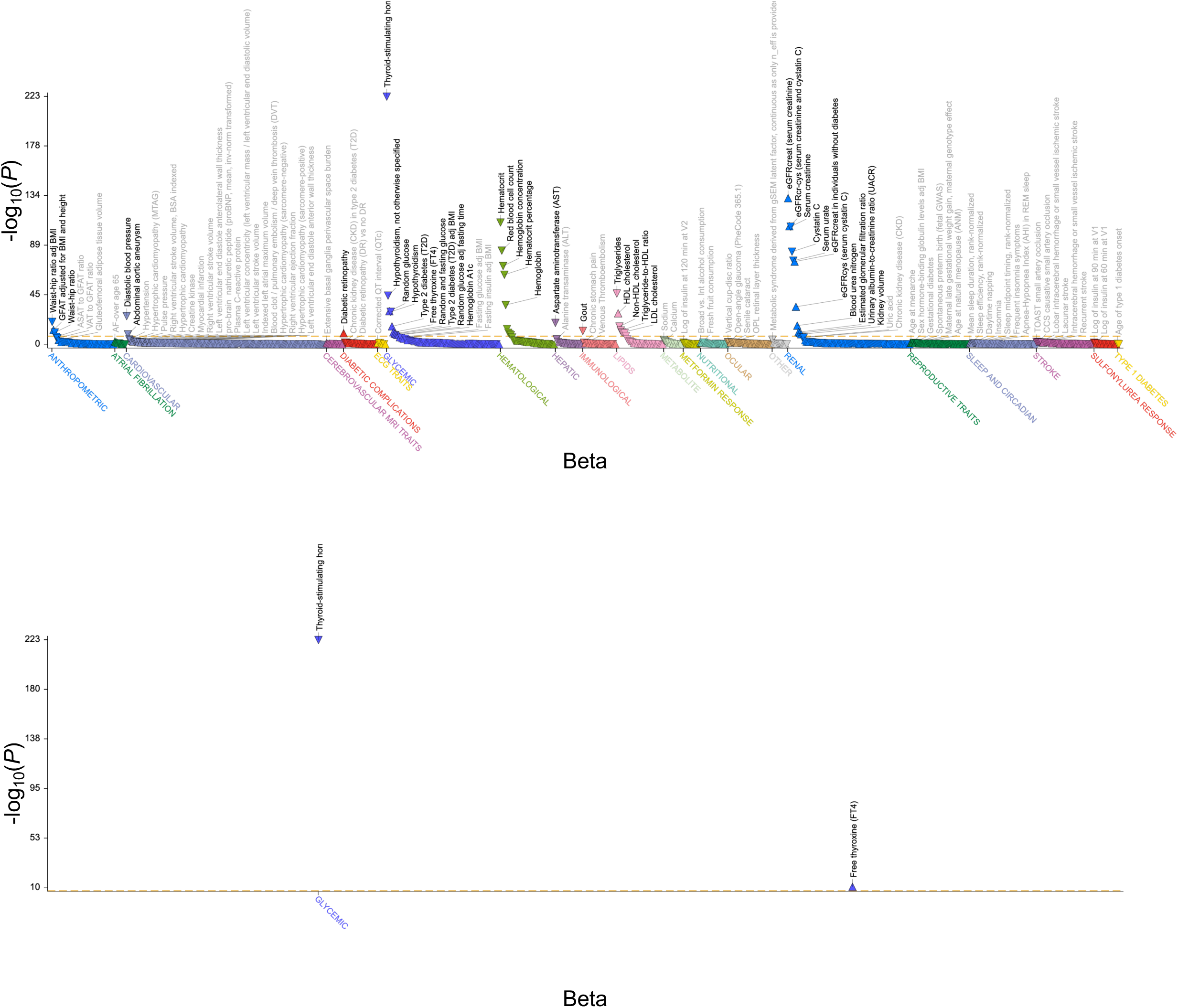
Phenome-wide association plot for rs9472135. Phenome-wide association results of rs9472135 using data from the Common Metabolic Disease Knowledge Portal (https://hugeamp.org) is shown on the top panel. The bottom panel shows a focused view displaying only the associations of rs9472135 with thyroid-stimulating hormone (TSH) and free thyroxine (FT4), plotted on the same scales, highlighting the specific effects of this variant on thyroid function traits. Each point represents a single phenotype, with the x-axis showing the effect size (β) and the y-axis showing the corresponding -log10(*P*) value for SNP–trait associations across the phenome.

**Supplementary Figure 9.**
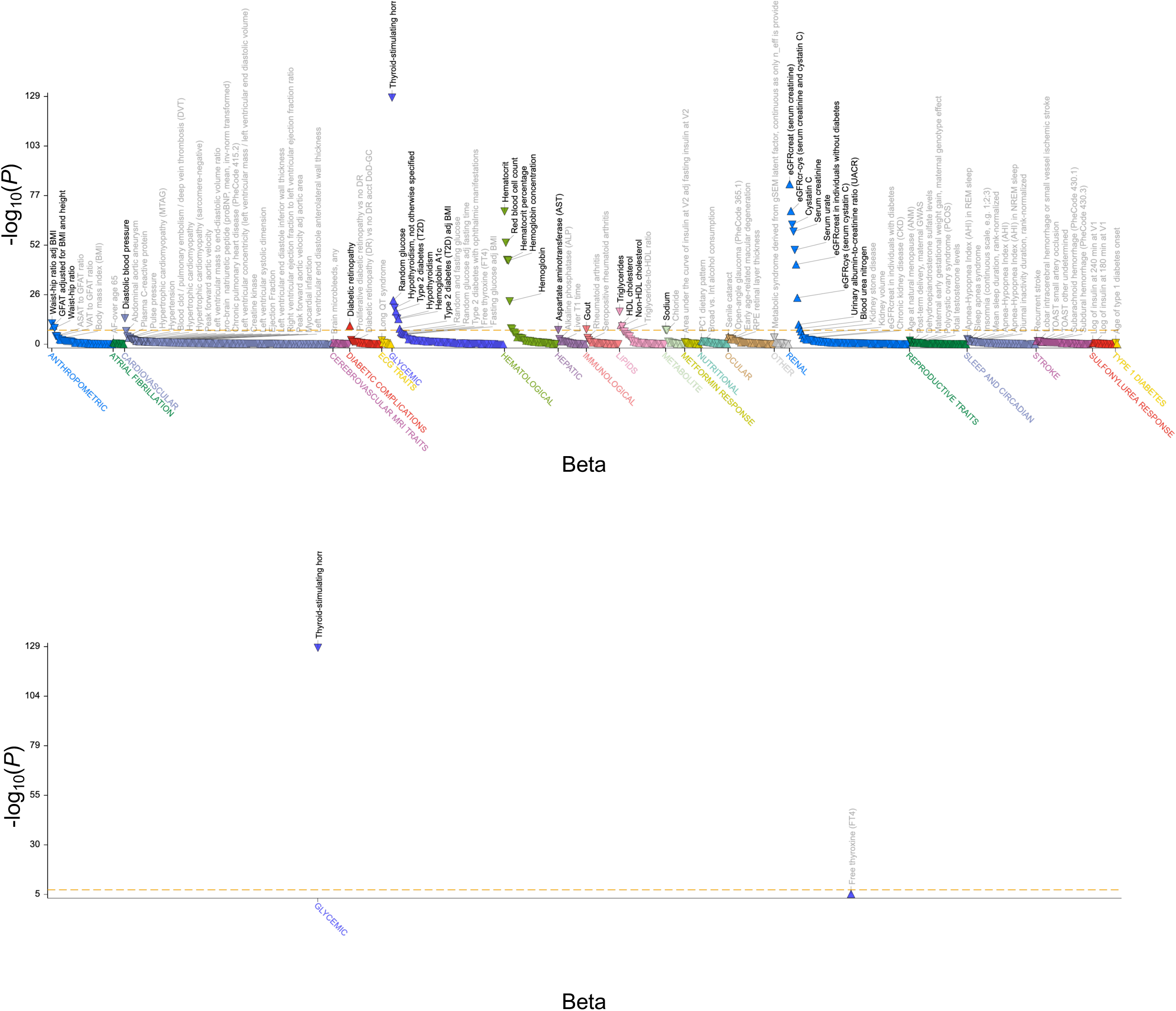
Phenome-wide association plot for rs4714704. Phenome-wide association results of rs4714704 using data from the Common Metabolic Disease Knowledge Portal (https://hugeamp.org) is shown on the top panel. The bottom panel shows a focused view displaying only the associations of rs4714704 with thyroid-stimulating hormone (TSH) and free thyroxine (FT4), plotted on the same scales, highlighting the specific effects of this variant on thyroid function traits. Each point represents a single phenotype, with the x-axis showing the effect size (β) and the y-axis showing the corresponding -log10(*P*) value for SNP–trait associations across the phenome.

**Supplementary Figure 10.**
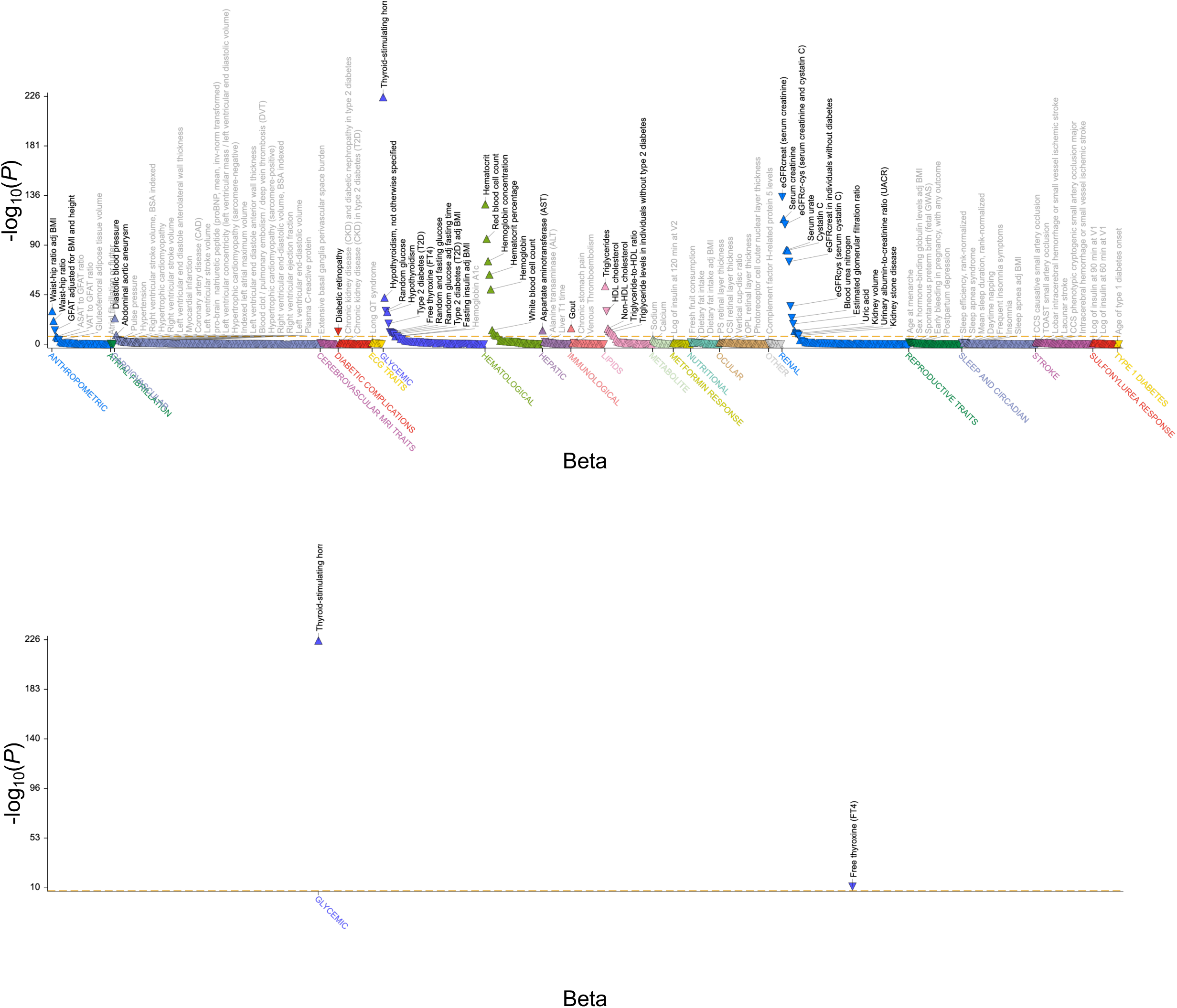
Phenome-wide association plot for rs10223666. Phenome-wide association results of rs10223666 using data from the Common Metabolic Disease Knowledge Portal (https://hugeamp.org) is shown on the top panel. The bottom panel shows a focused view displaying only the associations of rs10223666 with thyroid-stimulating hormone (TSH) and free thyroxine (FT4), plotted on the same scales, highlighting the specific effects of this variant on thyroid function traits. Each point represents a single phenotype, with the x-axis showing the effect size (β) and the y-axis showing the corresponding -log10(*P*) value for SNP–trait associations across the phenome.

**Supplementary Figure 11.**
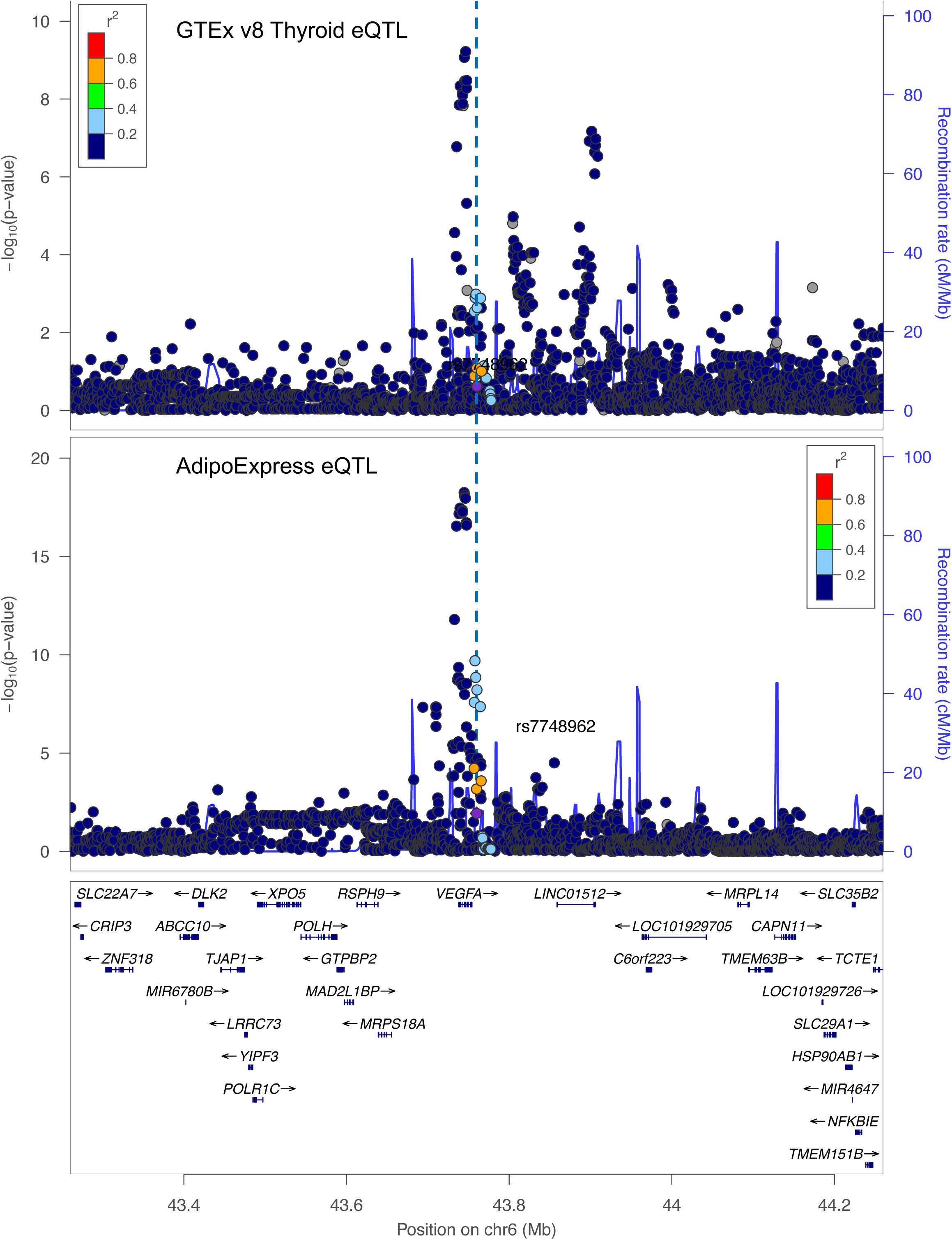
Regional *cis*-eQTL plot for rs7748962 in thyroid and adipose tissue. Regional *cis*-eQTL association plots for rs7748962 at the *VEGFA* locus. GTEx v8 thyroid *VEGFA* eQTL results are shown in the top panel, and AdipoExpress adipose eQTL results in the bottom panel. Variants across the locus are plotted by genomic position (x-axis) and - log10(*P*) (y-axis), with recombination rate superimposed and genes in the region, including *VEGFA* and neighboring loci, shown below.

**Supplementary Figure 12.**
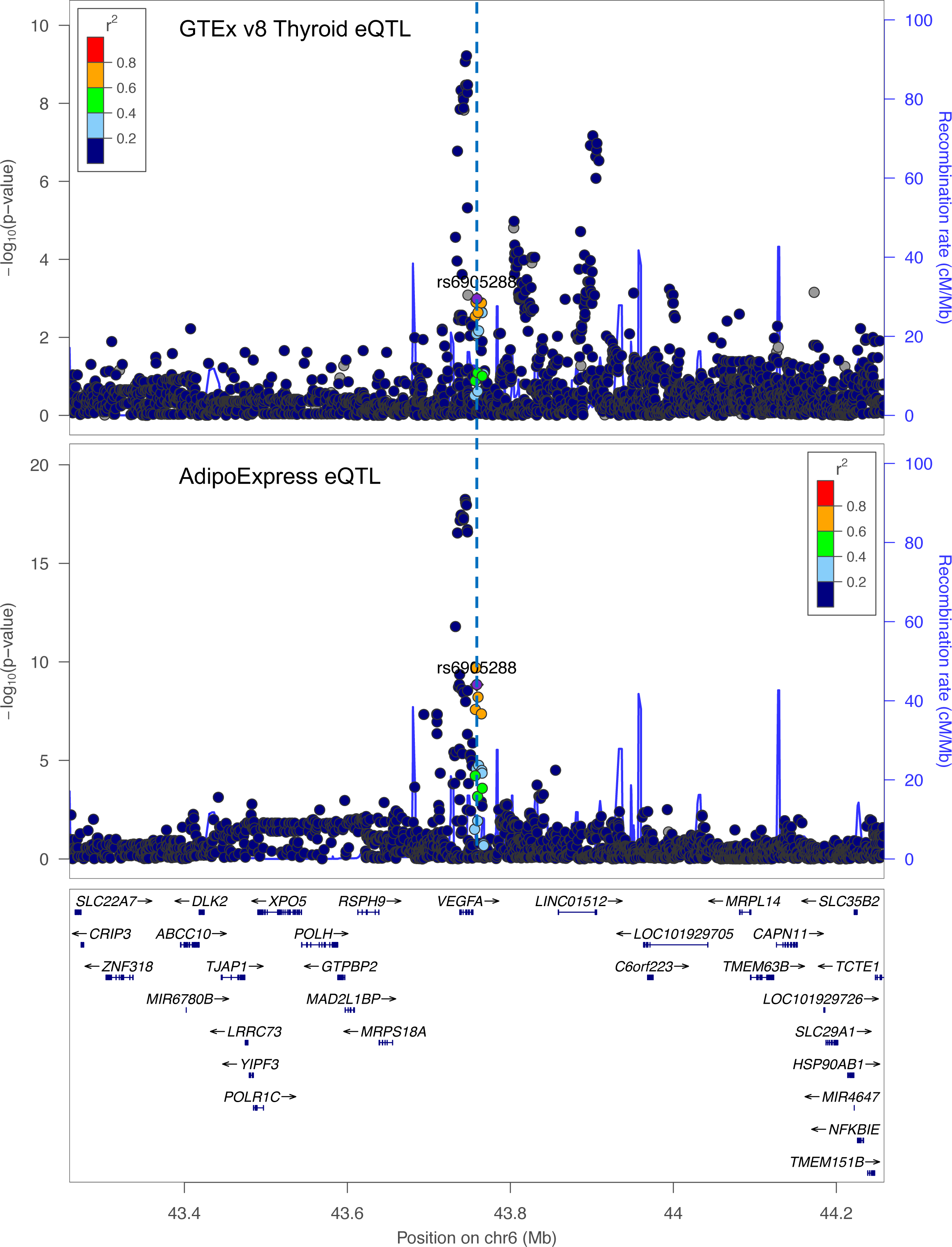
Regional *cis*-eQTL plot for rs6905288 in thyroid and adipose tissue. Regional *cis*-eQTL association plots for rs6905288 at the *VEGFA* locus. GTEx v8 thyroid *VEGFA* eQTL results are shown in the top panel, and AdipoExpress adipose eQTL results in the bottom panel. Variants across the locus are plotted by genomic position (x-axis) and - log10(*P*) (y-axis), with recombination rate superimposed and genes in the region, including *VEGFA* and neighboring loci, shown below.

**Supplementary Figure 13.**
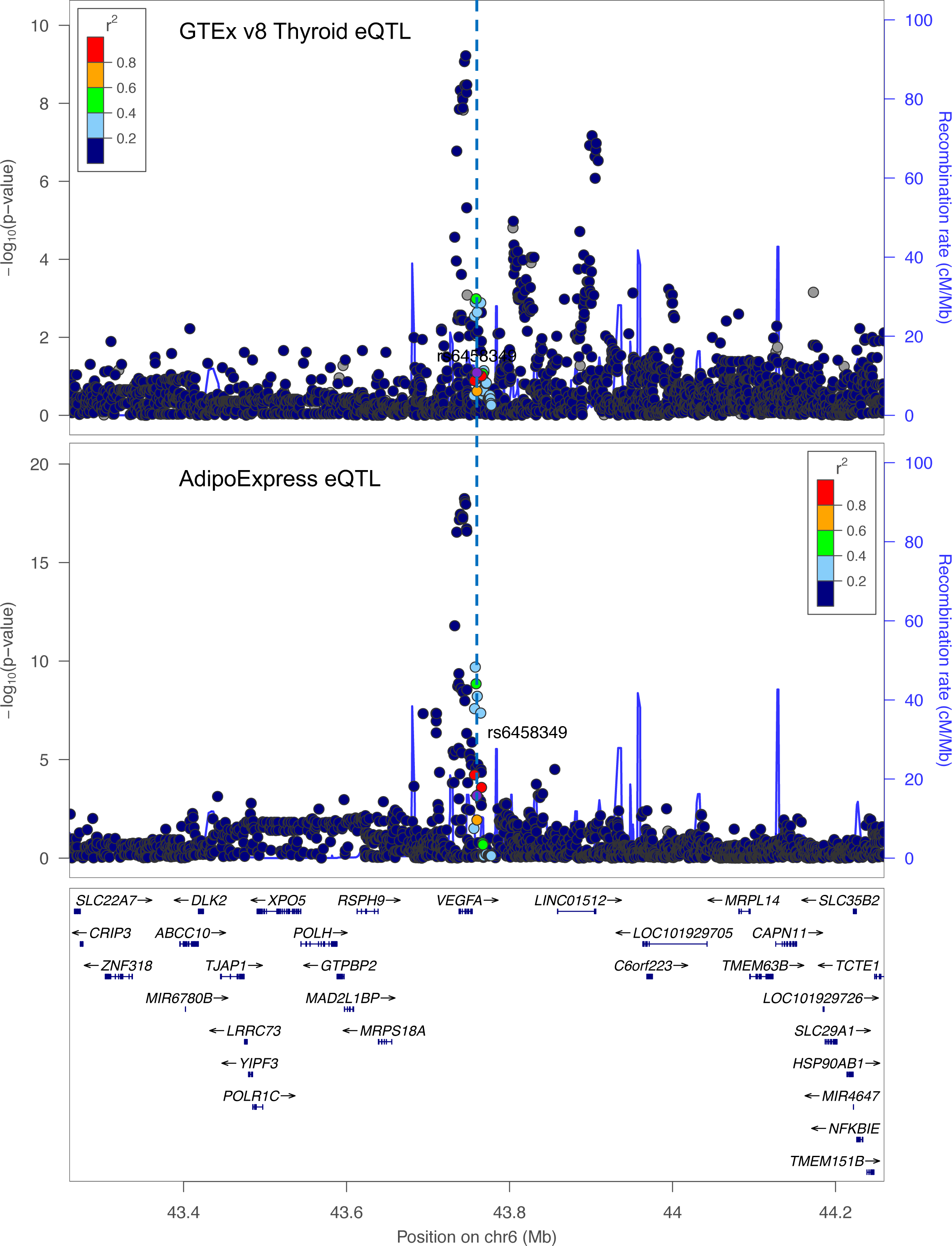
Regional *cis*-eQTL plot for rs6458349 in thyroid and adipose tissue. Regional *cis*-eQTL association plots for rs6458349 at the *VEGFA* locus. GTEx v8 thyroid *VEGFA* eQTL results are shown in the top panel, and AdipoExpress adipose eQTL results in the bottom panel. Variants across the locus are plotted by genomic position (x-axis) and - log10(*P*) (y-axis), with recombination rate superimposed and genes in the region, including *VEGFA* and neighboring loci, shown below.

**Supplementary Figure 14.**
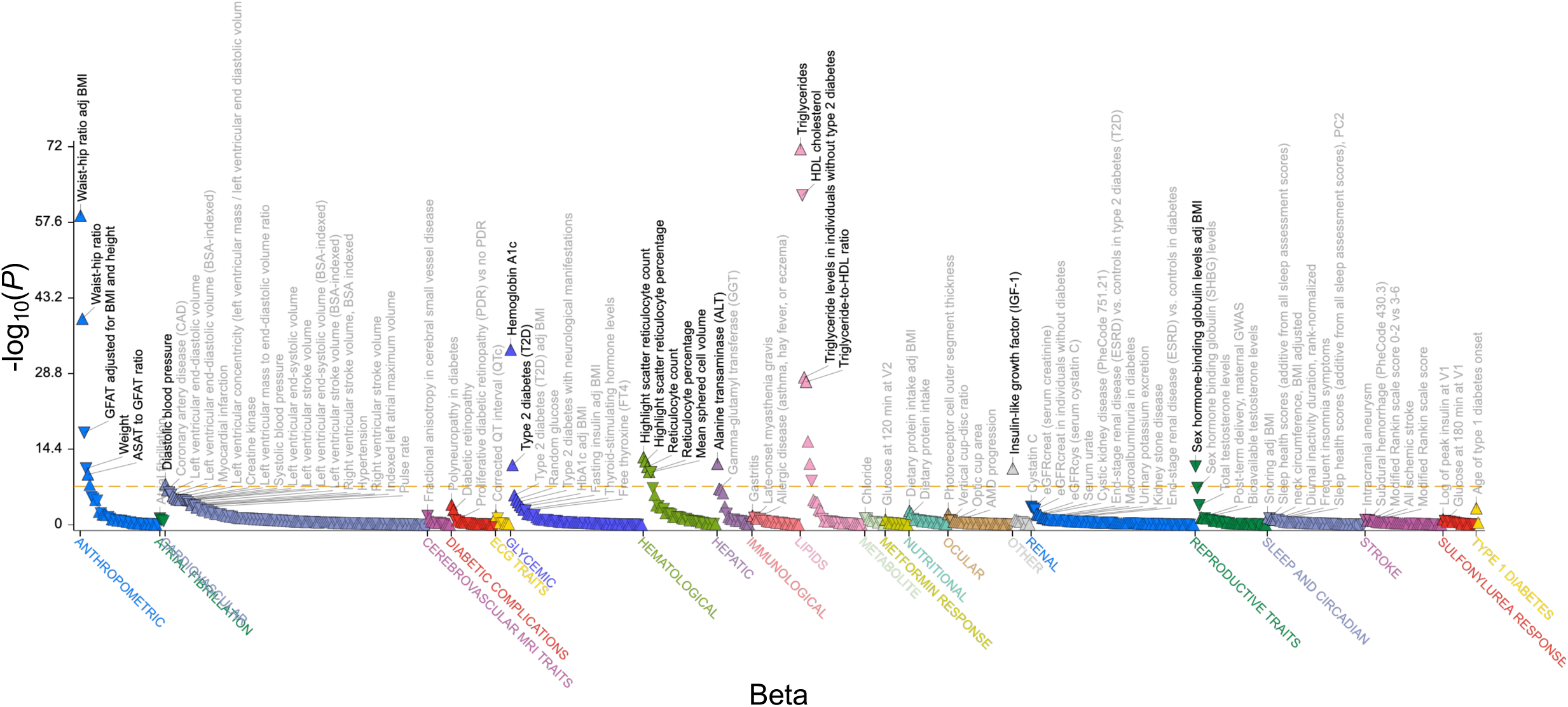
Phenome-wide association plot for rs7748962. Phenome-wide association results of rs7748962 (chr6:43,792,190-A) using data from the Common Metabolic Disease Knowledge Portal (https://hugeamp.org). Each point represents a single phenotype, with the x-axis showing the effect size (β) and the y-axis showing the corresponding -log10(*P*) value for SNP–trait associations across the phenome.

**Supplementary Figure 15.**
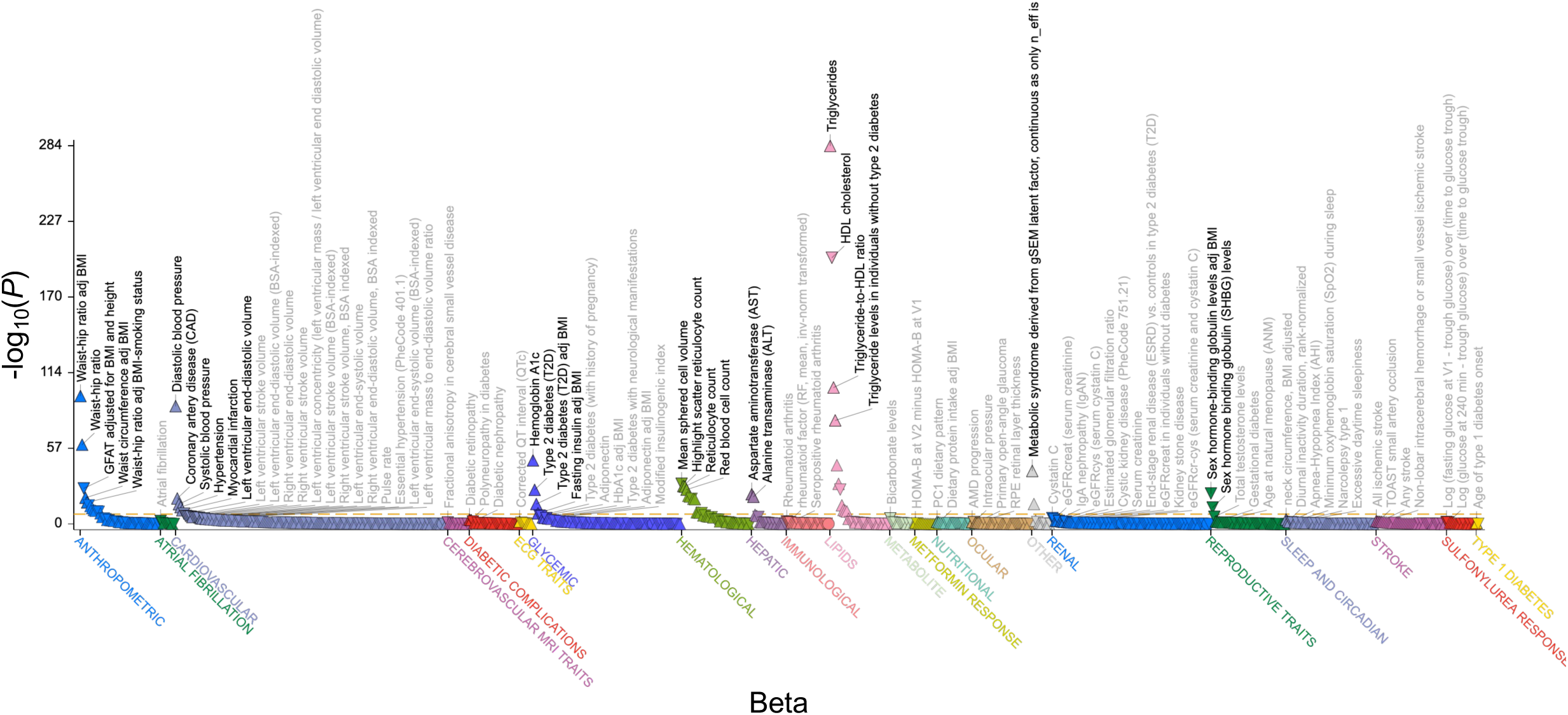
Phenome-wide association plot for rs6905288. Phenome-wide association results of rs6905288 (chr6:43,791,136-A) using data from the Common Metabolic Disease Knowledge Portal (https://hugeamp.org). Each point represents a single phenotype, with the x-axis showing the effect size (β) and the y-axis showing the corresponding -log10(*P*) value for SNP–trait associations across the phenome.

**Supplementary Figure 16.**
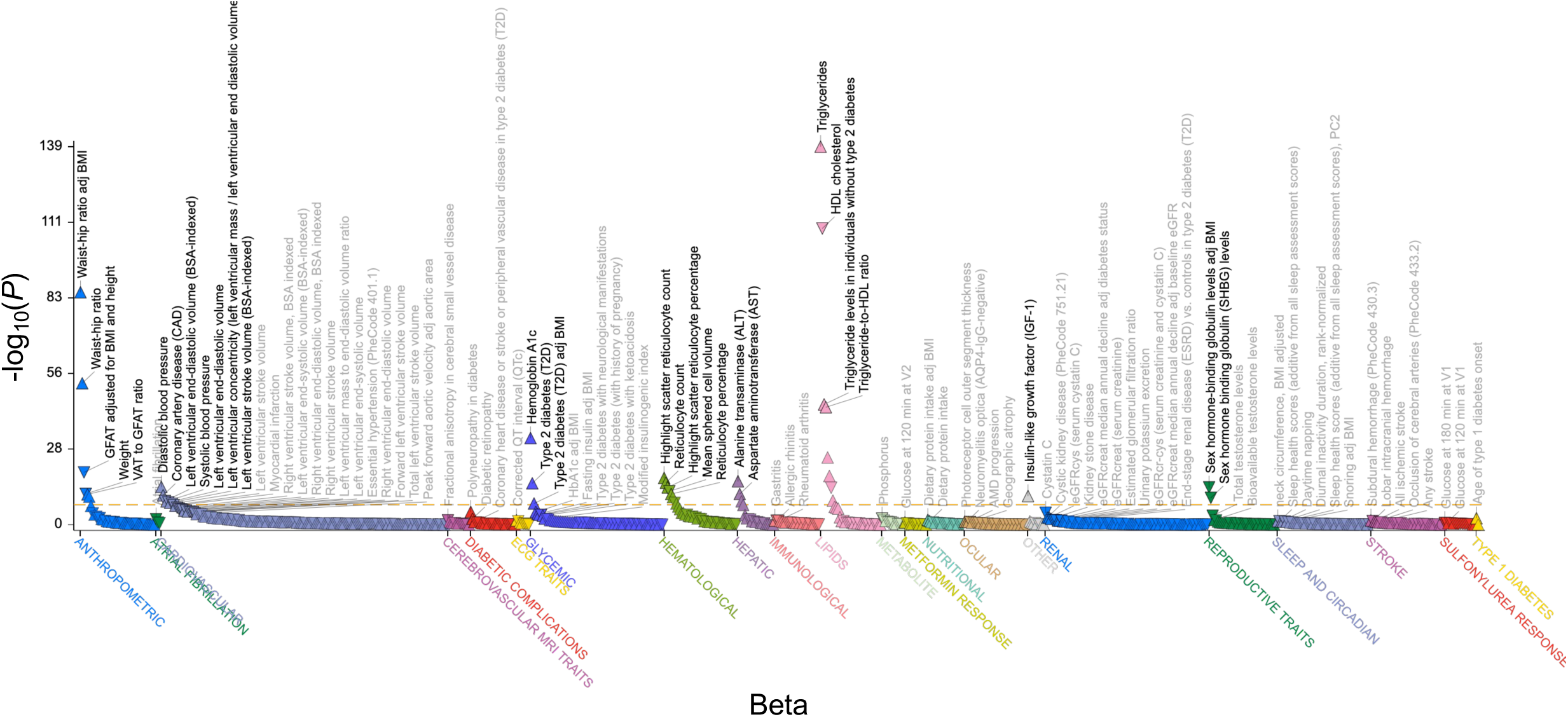
Phenome-wide association plot for rs6458349. Phenome-wide association results of rs6458349 (chr6:43,792,052-A) using data from the Common Metabolic Disease Knowledge Portal (https://hugeamp.org). Each point represents a single phenotype, with the x-axis showing the effect size (β) and the y-axis showing the corresponding -log10(*P*) value for SNP–trait associations across the phenome.

