## Supplementary Note for "Systematic variant-to-gene mapping highlights *TGFB2* and *VEGFA* as adipokine-coding genes with non-obese, insulin-resistance-like characteristics and distinct disease risks"

**Supplementary Note 1: Description of Million Veteran Program GWAS data**

**We used publicly available outcome GWAS for European, African, and East Asian ancestries from the Million Veteran Program (MVP) cohort which consists of up to 2,068 traits**^1^**. Participants in the MVP consist of a diverse range of United States Veterans who were grouped based on genetic similarity to the 1000 Genomes Project**^2^ **into population groups. Specifically, we assessed a total of 1,669 traits (1,470 binary, 88.1%; and 199 quantitative, 11.9%) in European ancestries (*n* = 449,042), 1,368 traits (1,187 binary, 86.8%; and 181 quantitative, 13.2%) in African ancestries (*n* = 121,177), and 241 traits (148 binary, 61.4%; and 93 quantitative, 38.6%) in East Asian ancestries (*n* = 6,702) (Supplementary Table 1, 2, and 3). Assessing the overlap of traits across ancestries, we found that 241 traits were shared among all three ancestries (Figure 1), 1368 between European and African, 241 between European and East Asian, and 241 between African and East Asian ancestries.**

**Supplementary Note 2: Multi-ancestry fine-mapping across the phenome in three populations**

**We identified 45,974 non-overlapping loci across 753 traits (33,179 loci shared between all three ancestries and 12,795 loci** shared between **European and African ancestries) for multi-ancestry fine-mapping (Supplementary Note Table 1 and Methods).**

**Upon cross-ancestry fine-mapping, we annotated each of the credible sets from each loci across all traits with a gene annotation provided by cS2G.** For each credible set, we grouped variants by linked gene and summed the posterior inclusion probabilities (PIPs) of all variants linked to that gene. Because cS2G can link one variant to multiple genes, a given variant could contribute to more than one gene; however, we did not additionally weight variant contributions by the cS2G linking score in this aggregation step. We then assigned the gene with the largest summed PIP as the representative effector gene for that credible set (**Methods**). **In total, we found 20,442 fine-mapped gene-trait pairs (PIP > 0.9), and 72,673 credible set-trait pairs (median CS size = 2) (Supplementary Note Figure 1). Previously, Kanai et al.**^3^ **performed single ancestry fine-mapping in Biobank Japan, FinnGen, and UK Biobank and found median credible set sizes of 11, 9, and 12, respectively. The substantial reduction in median credible set size when combining up to three ancestries in MVP demonstrates the increased resolution in pinpointing causal variants offered by including genetically diverse populations.**

Including genetically diverse ancestries for fine-mapping allows leveraging differences in linkage disequilibrium across genetic ancestries to improve fine-mapping. Earlier studies have shown that limited benefits are gained when combining European ancestries together or when combining European and East Asian ancestries^4,5^ while simulations show that including African ancestry may substantially reduce the size of credible sets^5^ likely due to their much narrower LD segments. Indeed, by leveraging 121,177 African ancestry MVP participants, our analyses support these arguments showing a median credible set size of 2 when cross-ancestry fine-mapping is performed across hundreds of traits.

Because external reference panels can exhibit LD patterns that differ from those in the discovery GWAS samples, relying on them may introduce bias into fine-mapping analyses^6^. To address this, we evaluated the LD consistency between MVP traits and reference samples for each fine-mapped loci in European, African, and East Asian ancestries by estimating the s value in the SuSiE-RSS model using regularized LD^7^. Across fine-mapped loci, 84.4%, 91.4%, and 97.4% in European, African, and East Asian ancestries had s values below 0.1, with median s values of 0.031, 0.016, and 0.0079, respectively, suggesting that summary statistics and reference panels had reasonably consistent LD. Further, in order to focus on shared credible sets and not ancestry-specific credible sets, we assessed the post hoc probability to ensure that fine-mapping was not predominated by a single ancestry such as European ancestries which had the largest sample size. In European, African, and East Asian ancestries, 93.1%, 34.9%, and 4.2% of fine-mapped loci had post hoc probabilities above 0.8, respectively (**Supplementary Note 3)**. Signals with post-hoc probability for the credible set in a population that surpassed 0.8 were considered causal in that population, as done previously^8^. All cS2G and fine-mapped loci with s value and post hoc probabilities are provided in **Supplementary Note Table 2**.

***Validation of integrative SNP-to-gene and multi-ancestry fine-mapping for prioritizing causal variants***

To verify the validity of combining variant-to-gene mapping and multi-ancestry fine-mapping to pinpoint causal genes, we tested the gene-trait pairs for overlap with a set of ExWAS-implicated genes for seven traits used previously^9^ (**Supplementary Note Table 3**). We used 7 publicly available traits with large number of cases or sample size in the UK Biobank pertaining to calcium level, standing height, hypothyroidism, low density lipoprotein cholesterol level, red blood cell count, triglyceride level, and T2D. The overlap ranged between 20.6% for red blood cell count to 54.5% for calcium levels (**Supplementary Note Table 3**). Our results indicate that cS2G and fine-mapping together can capture GWAS-relevant genes in specific traits.

Method details: Enrichment to validate cS2G and finemapping

To validate that combining cS2G and fine-mapping captures relevant genes, we used the same approach as a recent effector gene prioritization method by Schipper et al.^9^ who considered nine traits from Liang et al.^10^. We were able to assess seven of the nine traits which were present in MVP which included calcium level, standing height, hypothyroidism, low density lipoprotein cholesterol level, red blood cell count, triglyceride level, and T2D. Traits were downloaded from the AstraZeneca PheWAS Portal using the latest release “UK Biobank 500k WGS (v2) Public” (<https://azphewas.com/>).

For the AZPheWAS traits, we subsetted to EUR ancestry hits with P < 5e-8. For the cS2G and fine-mapping trait, we subsetted to credible sets which “passed in one ancestry”. The overlap was quantified by considering how many genes from the cS2G and finemapping results were present in the AZPheWAS genes.

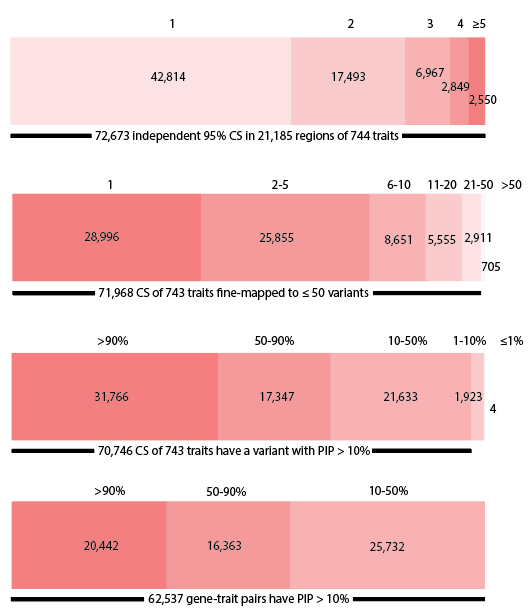

**Supplementary Note Figure 1**. **Multi-ancestry fine-mapping result summary.**

The number of independent 95% credible sets (CS) per locus, number of fine-mapped variants per 95% CS, number of 95% CS binned by the best PIP variant in each CS, and number gene-trait pairs binned by PIP after each CS has been assigned a gene using cS2G.

**Supplementary Note 3: Post-hoc probability statistics**

Multi-ancestry fine-mapping resulted in the identification of 72,673 total credible sets composed of 744 traits and 21,185 loci. In European (EUR), African (AFR), and East Asian (EAS) ancestries, we identified 67,654, 25,374, and 2,212 credible sets with post-hoc probability > 0.8, respectively.

We also found that 20,825 credible sets had post-hoc probability > 0.8 in both EUR and AFR pertaining to 515 unique traits and 8,992 loci.

We observed 1,193 credible sets with post-hoc probability > 0.8 in EUR and AFR and EAS which included 121 unique traits and 707 loci.

**Supplementary Note 4: Four *VEGFA* variants in Thyroid-Adiposity cluster are associated with expression changes in long intergenic non-protein coding RNA**

**Interestingly, these** four *VEGFA* variants **were also all significantly associated with expression changes in** long intergenic non-protein coding RNA 02537, ***LINC02537* (*P* < 1.8e-17) in thyroid tissue and aside from rs4714704, the other three variants were also *LINC01512* eQTLs in thyroid (*P* < 4.4e-6). Moreover, we observed that the four variants were all associated with alternative splicing patterns of *LINC02537* in the thyroid (*P* < 1.9e-7) and testis (*P* < 9.8e-6) suggesting that these variants may have broader regulatory roles affecting both gene expression and splicing, potentially influencing the transcriptional landscape of multiple genes in a tissue-specific manner (Supplementary Table 9). This highlights the complexity of variant-driven regulation at the *VEGFA* locus, particularly in tissues like thyroid and testis where coordinated regulation of neighboring non-coding RNAs may play a functional role.**

**Supplementary Note 5: *VEGFA* region demonstrates pleiotropy**

**In the *VEGFA* region, we observed** multiple clear peaks in thyroid eQTL whereas adipose tissue was dominated by a single peak (**Supplementary Figure** **3-6** and **Supplementary Figure 14-16**). The three GTEx thyroid peaks for *VEGFA* are located in the gene regions for *VEGFA* (lead *cis*-eQTL: rs3025000; 6:43746169_C/T), *LINC02537* (lead *cis*-eQTL: rs729761; 6:43804571_T/G) and *LINC01512* (lead *cis*-eQTL: rs6923866; 6:43901184_T/C), respectively. Interestingly, all four *VEGFA* variants from Cluster 2 (thyroid-adipose cluster) were located in the middle peak within the *LINC02537* gene region whereas all three VEGFA variants from Cluster 3 (Lipodystrophy cluster) were found in the leftmost peak within the *VEGFA* gene region. Further, the lead *cis*-eQTLs in these gene regions showed interesting profiles. While rs3025000 in the *VEGFA* loci was an eQTL for *VEGFA* in thyroid, rs729761 in *LINC02537* was eQTL for both *VEGFA* and *LINC02537* and the peak in *LINC01512*, rs6923866, was eQTL for both *VEGFA* and *LINC01512*. Additionally, all three lead *cis*-eQTLs had eQTL evidence for *VEGFA* in adipose tissue from AdipoExpress where the *VEGFA* variant, rs3025000, had the strongest association as expected (*P* = 1.1e-18) (**Supplementary Table 9**).

The lead variants in *LINC02537* (rs729761) and *LINC01512* (rs6923866) loci did not have eQTL in adipose tissue at chromosome 6 for their respective LINC genes since AdipoExpress did not include *LINC02537* or *LINC01512*. Corroborating this, based on GTEx v8, the expression levels of these two LINC RNA appeared to be low in adipose tissues. Hence, this suggests that no eQTL was found in AdipoExpress for *LINC02537* and *LINC01512* potentially due to lack of detection of these two transcripts.

In summary, we pinpointed Cluster 3 variants as predominantly adipose-specific regulators of *VEGFA*, while Cluster 2 variants demonstrated dual regulatory activity on both *VEGFA* and *LINC02537* in thyroid tissue, indicating a pleiotropic mechanism with potentially divergent biological consequences (see **Supplementary Note 5: Discussion** below).

***Supplementary Note 5: Discussion***

Two distinct genetic clusters near *VEGFA* appear to exert regulatory effects on this gene, yet their tissue specificity and downstream targets differ. One cluster seems to act primarily in adipose tissue, targeting *VEGFA* alone, whereas the other cluster exhibits a more complex regulatory profile, influencing both *VEGFA* and *LINC02537* in thyroid tissue with opposing directions of effect. This pattern suggests that while the regulatory impact on *VEGFA* itself is consistent, the presence of pleiotropic effects involving *LINC02537*—whose function remains poorly understood—may drive the separation of these clusters. These findings support the notion that although these variants were mapped to *VEGFA*, there may be distinct signals acting through *VEGFA*.
